# Randomized controlled trial of gut decontamination in pediatric patients undergoing allogeneic hematopoietic cell transplantation

**DOI:** 10.1101/2021.12.16.21267940

**Authors:** Christopher J. Severyn, Benjamin A. Siranosian, Sandra Tian-Jiao Kong, Angel Moreno, Michelle M. Li, Nan Chen, Christine N. Duncan, Steven P. Margossian, Leslie E. Lehmann, Shan Sun, Tessa M. Andermann, Olga Birbrayer, Sophie Silverstein, Soomin Kim, Niaz Banaei, Jerome Ritz, Anthony A. Fodor, Wendy B. London, Ami S. Bhatt, Jennifer S. Whangbo

## Abstract

**Background:** Gut decontamination (GD) can decrease the incidence and severity of acute graft-versus-host-disease (aGVHD) in murine models of allogeneic hematopoietic cell transplantation (HCT). Several HCT centers standardly practice GD with different antibiotic regimens. In this pilot study, we examined the impact of GD on the gut microbiome composition and incidence of aGVHD in HCT patients.

**Methods:** We randomized 20 pediatric patients undergoing allogeneic HCT to receive (GD) or not receive (no-GD) oral vancomycin-polymyxin B from day -5 through neutrophil engraftment. We evaluated shotgun metagenomic sequencing of serial stool samples to compare the composition and diversity of the gut microbiome between study arms. We assessed clinical outcomes in the 2 arms and performed strain-specific analyses of pathogens that caused bloodstream infections (BSI).

**Results:** The two arms did not differ in Shannon diversity of the gut microbiota at two weeks post-HCT (Genus, *p=*0.8; Species, *p=*0.44) or aGVHD incidence (*p*=0.58). Immune reconstitution of T-cell subsets was similar, but absolute CD19+ B-cell counts were higher in the GD arm at 12 months post-HCT (*p*=0.02). Five patients in the no-GD arm had eight BSI episodes vs one episode in the GD arm (*p*=0.09). The BSI-causing pathogens were traceable to the gut in seven of eight BSI episodes in the no-GD arm, including the genus *Staphylococcus*.

**Conclusions:** While GD did not differentially impact Shannon diversity or clinical outcomes, our findings suggest that GD may protect against gut-derived BSI in HCT patients by decreasing the prevalence or abundance of gut microbial pathogens.

**Key points:**

– In this phase 2 randomized study of gut decontamination (GD) in 20 pediatric HCT patients, neither two-week post-HCT Shannon diversity of the gut microbiome nor incidence of aGVHD differ between the GD and no-GD arms.
– All bloodstream infections (BSIs) caused by pathogens traceable to the gut either temporally or via strain-specific analysis (concomitant gut colonization) occurred in patients in the no-GD arm; this suggests that GD with vancomycin-polymyxin B may decrease the incidence of gut-derived BSI in allo-HCT patients.
– In contrast to prior studies, we find that non-mucosal barrier injury (MBI) pathogens, such as *Staphylococcus aureus,* can be found in the gut microbiome of HCT patients.

## INTRODUCTION

Despite advances in graft manipulation and acute graft-versus-host-disease (aGVHD)- prophylactic regimens, aGVHD and other treatment related complications remain major causes of morbidity and mortality in patients undergoing allogeneic hematopoietic cell transplantation (allo-HCT). In efforts to decrease the incidence and severity of aGVHD, gut decontamination (GD) with non-absorbable antibiotics in the peri-HCT time-period is administered in some clinical centers, largely based on limited preclinical studies. Specifically, mouse and human studies in the 1970s and 1980s demonstrated that debulking or eradication of intestinal bacteria by exposure to non-absorbable oral antibiotics could decrease the risk of aGVHD [1–4]. A retrospective study of patients transplanted from 1989 to 2002 (n=112) reported that subjects who were successfully decontaminated, as defined by stool culture after exposure to GD, had significantly lower rates of aGVHD compared to those without successful GD [5]. While the precise mechanism of how GD impacts aGVHD remains unknown, it is thought that decreasing the total microbial load or creating a specific balance of microbes can improve intestinal barrier integrity and decrease inflammation via interactions with the host immune system (reviewed in [6, 7]).

In addition to non-absorbable antibiotics, systemic antimicrobial prophylaxis has also been used with the intent of suppressing bacterial growth in the gut. For example, a prospective study by Beelen *et al*. showed that patients randomized to receive ciprofloxacin plus metronidazole had a lower incidence of grade II-IV aGVHD compared to those randomized to receive ciprofloxacin alone (25% vs 50%, p<0.01) [8]. In this study, prophylaxis with ciprofloxacin plus metronidazole was associated with a lower amount of culturable anaerobic bacteria in the gut microbiome, suggesting a link between anaerobic bacteria and aGVHD. However, other studies have shown that the use of systemic antibiotics with activity against anaerobic bacteria is associated with an increased risk of aGVHD [9–11]. If the effect of GD on the pathogenesis of aGVHD is related to the types of gut microbes that are cleared and the extent to which they are eradicated, then measuring the impact of GD on the various organisms that comprise the microbiome may help to resolve these discordant findings.

There are many limitations to prior clinical trials for GD. First, comparisons between trials are confounded by the variations in the practice of GD and systemic prophylaxis across centers with no consensus among the choice of antibiotic regimen (reviewed in [7]). In addition, many GD studies are single-arm trials without a comparator arm, or when a comparator arm is included there is lack of a no-GD arm, instead comparing two different GD interventions to one another. As noted above, successful GD as measured by culture-based approaches was associated with improved outcomes. However, a limitation of these culture-based approaches is that many organisms within the microbiome, such as strict anaerobes, are difficult to culture in samples that are not immediately stored in anaerobic conditions [7]. Modern next generation sequencing (NGS)-based approaches enable a more comprehensive profiling of the gut microbiome composition by overcoming the need to maintain viability of the organisms within the stool sample, and thus may help inform the specific impact of GD on the gut microbiome. Indeed, a recent study using NGS demonstrated that low microbial diversity in the stool at the time of neutrophil engraftment after allo-HCT is associated with decreased survival [12]. Despite the above studies, it remains unclear whether GD is beneficial for lowering the risk of aGVHD in HCT patients.

While aGVHD is the most prevalent and fatal treatment related complication of HCT, bloodstream infections (BSI) are also an important cause of treatment-related morbidity and mortality in patients undergoing allo-HCT. The cumulative incidence of BSI in pediatric HCT patients is approximately 20% in the first 100 days [13] (ranging from 15% to 65% [13–17]), with 18% BSI- attributable mortality (range 12% to 20%) [13], and an estimated healthcare cost of $40,000 to $70,000 per BSI incident [14, 18]. In adult HCT patients, a diverse gut microbiome is associated with a lower risk of chemotherapy-related BSI during HCT [17]. Multiple studies have shown the microbiota composition may predict BSI in adult [17, 19] and pediatric HCT patients [20], and during chemotherapy for pediatric acute leukemia [21]. GD has been explored in the granulocytopenic population as a strategy to reduce BSI [22], and to a limited extent in HCT patients, with no difference compared to historic controls [23] nor comparing BSI in a hematology (no-GD) versus HCT ward (GD) [24]. GD has been much more extensively studied in other vulnerable patient populations. For example, the largest cohort of patients to receive GD with the intent to decrease the risk of BSI has been patients in intensive care units (ICU), regardless of underlying diagnosis. These ICU studies have reported conflicting results, with some studies showing a reduction in BSI [25], while others report no significant differences in Gram-negative bacteremia [26].

BSIs in HCT patients are the consequence of infectious pathogens entering the bloodstream through indwelling catheters, breakdown in the skin, and mucosal barrier injury (MBI) secondary to conditioning chemotherapy and resulting neutropenia. Changes in the abundance of MBI- associated enteric strains often precede BSI episodes in patients undergoing HCT [17, 20, 27]. In addition to MBI, different bacterial strains of the same species behave differently [28], leading to a more prevalent strain under a selective pressure [29, 30]. Being able to identify and subsequently analyze strain variation [31] could aid in a better mechanistic understanding of how BSIs occur in patients.

We carried out the first prospective, randomized study of GD using oral, non-absorbable antibiotics (vancomycin and polymyxin B) to assess the impact of GD on the gut microbiome composition and diversity, and secondary outcomes such as the incidence of aGVHD and immune reconstitution. Based on recent studies [9–12], we hypothesized that GD would lead to decreased diversity of the gut microbiota, and potentially worse clinical outcomes. In an exploratory analysis, we investigated whether GD is associated with a decreased incidence of BSI. We observed that all BSI microbes that could be traced to the gut were from the no-GD arm, forming the basis for larger clinical and translational studies. We also used strain-specific analysis to demonstrate that the gut may be a reservoir for pathogens that have traditionally been classified as non-MBI pathogens, including *Staphylococcus aureus*.

## RESULTS

### Patient characteristics are similar between the two arms

Twenty pediatric patients undergoing allogeneic HCT were enrolled and randomized between March 2016 and June 2019 (Supplemental Figure 1). Ten patients received GD per our institution’s standard of care with oral non-absorbable vancomycin and polymyxin B from day -5 relative to the transplant through neutrophil engraftment (dosed according to body surface area in Supplemental Table 1, with actual administration in Supplemental Figure 2), and 10 patients received no GD (Figure 1). Baseline characteristics between the two arms were similar (Table 1). The median age at HCT for all patients was 15.2 years (range, 7.1-24.6). Most patients had underlying hematologic malignancies (n=15), received myeloablative conditioning regimens (n=17), and had bone marrow as the graft source (n=19). A median of seven stools per patient per transplant (range, 3-18) were collected starting pre-transplant through one-year post-transplant. 76% of the stool samples were collected within the first 30 days after HCT (Supplemental Figure 3).

**Figure 1.**
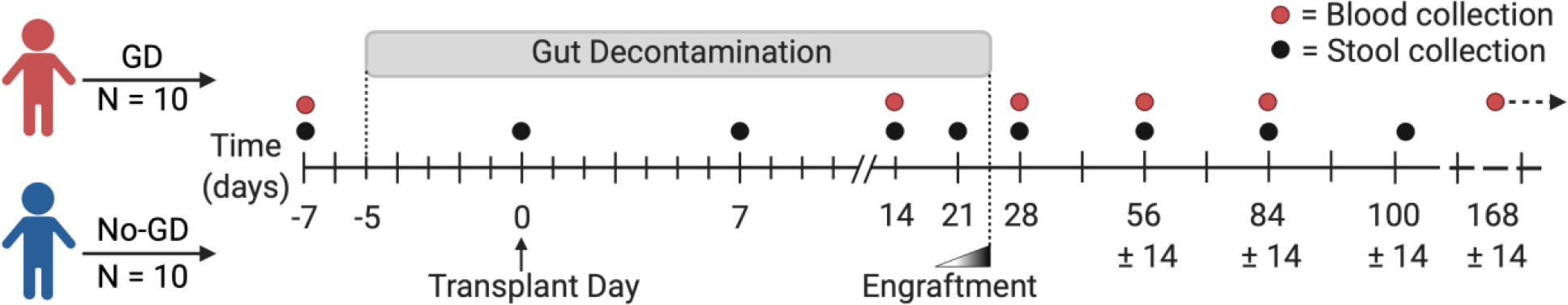
Study design. A total of 20 patients undergoing allo-HCT were randomized to two arms, 10 patients in the Gut Decontamination (GD with vancomycin-polymyxin B) and 10 patients in the no-GD arm. The GD arm received Vancomycin-polymyxin B, given three times daily, with dosing based on body surface area and analyzed as intention-to-treat (Supplemental Table 1). GD was given starting day -5 through neutrophil engraftment which is variable depending on the patient (median neutrophil engraftment day +25, see Supplemental Figure 2). The no-GD arm had the same stool and blood collection time points and did not receive oral Vancomycin-polymyxin B. Closed black circles are time of stool collections including pre-transplant, weekly until engraftment, and monthly until day +100. An additional cohort of two healthy sibling donors serve as a stool control comparison group (Supplemental Fig. 8). For immune reconstitution studies, blood samples (red circles) were collected at pre-transplant, 2 weeks, monthly for the first 3 months, then months 6, 9, 12, 18, and 24 (red circle with right arrow).

**Table 1.**
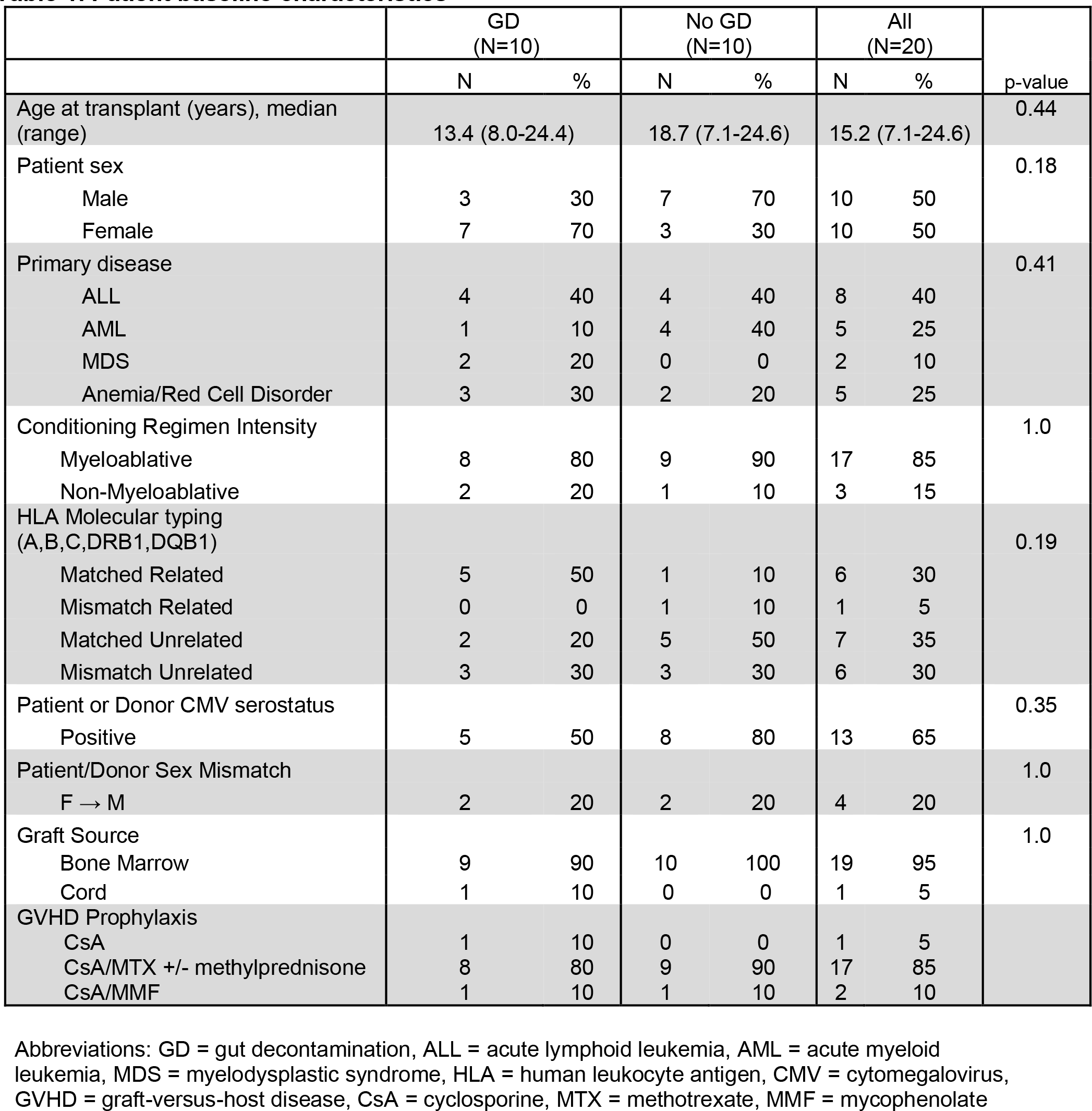
Patient baseline characteristics.

### Shannon diversity decreased similarly in patients with or without GD

To compare Shannon diversity within the gut microbiome between individuals on the two arms of the study, we first sought to determine the taxonomic composition of the gut microbiota using the time-series stool collections. DNA was extracted from 147 patient stool samples and 142 (of 147, 97%) were subjected to whole-genome shotgun (WGS) metagenomic library preparation and short-read sequencing (Supplemental Figure 3; the remaining 5 of 147 (3%) were unable to be sequenced due to insufficient biomass of the sample). Libraries were sequenced to a raw mean depth of 63.7x10^6^ read pairs (median 72.3x10^6^ read pairs; range 3.7x10^6^-338x10^6^ read pairs) per stool sample. After pre-processing and quality control filtering of raw reads, a median 16.9 x 10^6^ (range 4.3x10^4^-81x10^6^) high quality reads per sample were obtained for fecal metagenomes (Supplemental Figure 4). In most cases where a large proportion of reads was removed during pre-processing (n=2 with less than 1 x 10^5^ reads, n=14 of 142 (10%) with less than 1 x 10^6^ reads), this was due to a very high proportion of human (non-microbial) reads within the sample. Taxonomic composition was determined using Kraken2 classification [32] against a database of all bacterial, fungal, and viral genomes contained in NCBI Genbank as of January 2020.

To test the hypothesis that GD would lead to a significant decrease in diversity of the gut microbiota, we focused on the primary endpoint of Shannon diversity at 2 weeks post-HCT. At this selected time point, stool specimens likely reflect changes induced by the conditioning regimen and gut decontamination in the GD arm, are collected prior to the development of inflammation and acute GVHD [33], and precede the use of aGVHD-targeted immunosuppressive medications. Shannon diversity, which is a measurement sensitive to the loss of rare taxa [34] and estimates microbial richness (e.g., the number of species) and evenness (e.g., the relative abundance of organisms within a sample), was calculated for each sample (Supplemental Figure 5). Consistent with previous studies [12, 17, 35], the median Shannon diversity of the gut microbiota at the species level prior to GD exposure was 3.6 (range, 2.1-4.5) for GD and 3.3 (range, 1.2-4.2) for no-GD, and decreased at 2-weeks post-transplant to 2.4 (range, 0.03-5.32) for GD and 3.1 (range, 2.1-3.7) for no-GD (Figure 2A, Supplemental Table 3, Supplemental Figure 6A for genus). The two arms were similar in Shannon diversity at baseline (prior to GD exposure; Figure 2B for species (*p=*0.35), Supplemental Figure 6B for genus (*p=*0.32), Supplemental Table 4). At two-weeks post-transplant, there were no apparent differences in the Shannon diversity index between the GD versus no-GD arms for either species level (*p*=0.44, Figure 2B) or genus level (*p*=0.80, Supplemental Figure 6B). Furthermore, in an exploratory analysis, there was similar Shannon (alpha) diversity between the two arms when extending the window beyond 30 days to include all samples in the study (Supplemental Figure 7). We also calculated beta diversity using a pairwise Bray-Curtis dissimilarity index to determine how different the samples are from one another. When excluding samples from three patients (C05, C07, and C22) with >45% relative abundance of *Enterococcus faecium,* the beta diversity appeared similar between the two arms (Supplemental Figure 8). As a control, stool samples from two healthy sibling donors were also collected to serve as a comparison to the HCT patients, which were similar at the time points collected (based on analysis of similarity (ANOSIM), see gray circles and legend in Supplemental Fig. 8). Based on these findings, there is no evidence of a significant difference in our intention-to-treat analysis of the gut microbiota, per Shannon diversity, between the GD (n=10) versus no-GD (n=10).

**Figure 2.**
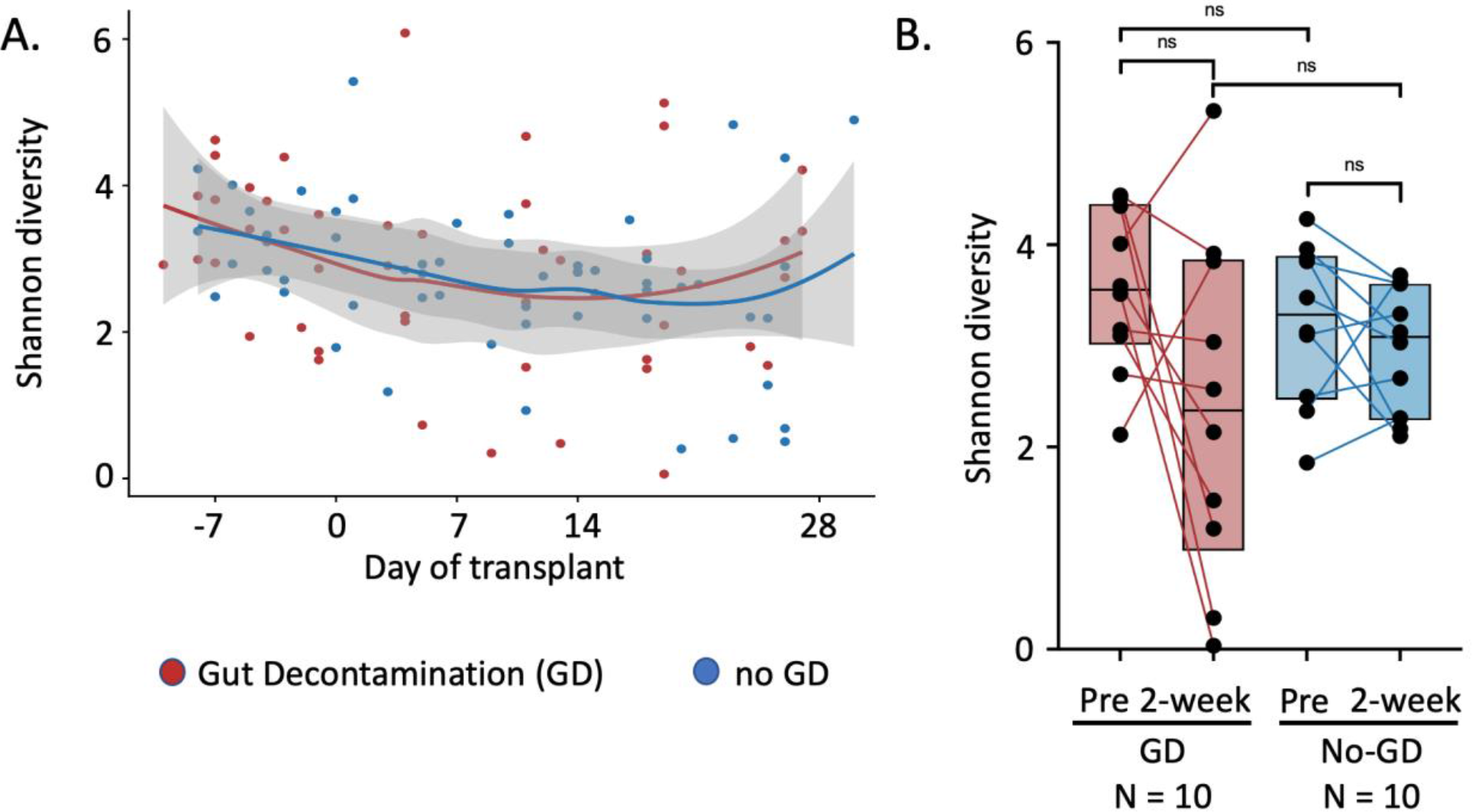
Shannon diversity is similar between the GD and no-GD groups based on intention-to-treat analysis at the species taxonomic level. Samples from patients undergoing GD (red) and no-GD (blue). (A) Shannon diversity over time analyzed at the species level using local polynomial regression fitting (LOESS-locally estimated scatterplot smoothing of the mean Shannon diversity) showing similarity between the two groups. (B) Shannon diversity of individual patients from pre-transplant (before GD antibiotics) to 2 weeks post HCT connected with a line. Boxes shown are the median with hinges at the 25% and 75%. All comparisons not significant (see Supplemental Table 4 for details). N=10 subjects GD arm, N=10 subjects no-GD arm. Comparison at the genus level (see Supplemental Figure 6 and Suppl. Tables 3 and 4) is also not significant (ns).

### No difference was observed in cumulative exposure to antibiotics between GD and no-GD arms

We sought to interrogate the reason that Shannon diversity within the gut microbiome did not differ between the two arms throughout the time course of evaluation. While there was varied adherence to vancomycin-polymyxin B (Supplemental Figure 2), the small sample size precludes a robust subset analysis comparing those in the GD arm who received a high proportion (>70% doses) of the planned GD (n= 4) versus those that had poor adherence (<30% doses, n=2). As GD might impact pathogen colonization in the gut microbiome and possibly subsequent bloodstream translocation of these pathogens, we postulated that GD might be associated with decreased fevers and thus decreased overall antibiotic exposure. In an exploratory analysis of the difference in systemic antibiotics between the two arms, we analyzed the clinical records based on individual antibiotics, class of antibiotics, and clinical indication including broad-spectrum antibiotics with anaerobic coverage (e.g., piperacillin-tazobactam, meropenem). We found no evidence of difference in the duration of prophylactic and therapeutic antibiotic exposure (within 30 days post-HCT) between the two treatment arms (Supplemental Table 5), including no evidence of a difference in broad-spectrum antibiotics with anaerobic coverage (median 13 days (GD) versus 17 days (no-GD), *p*=0.68). Thus, it is possible that the impact of GD was inconsequential compared to the impact of systemic broad-spectrum antibiotics.

### Comparison of secondary clinical outcomes

The pre-specified secondary outcomes of this study were stool frequency in the first 7 days, incidence of aGVHD in the first 100 days, relapse-free survival, overall survival (OS), and immune cell reconstitution. We explored the possibility of differences in the clinical outcomes of the patients undergoing GD compared to no-GD. Although antibiotic associated diarrhea can result from disruption of the gut microbiota, there was no apparent difference in the incidence of diarrhea in the first seven days post-transplant between the two treatment arms (Table 2*, p*=1.0). The overall incidence of grade 2-4 acute GVHD was 20%, with one patient in the GD arm and three patients in the no-GD arm (p=0.58, Table 2). The median day of onset of aGVHD was 38 days post-HCT (range 24 - 63). Of the 15 patients with a malignancy, three patients in the GD arm (n=7) and two patients in the no-GD arm (n=8) had malignant relapse within two years post-HCT; the 1-year relapse-free survival was 73±11.4% (n=15). The one-year OS was 100% (N=20, Supplemental Figure 9). No known harm from the GD treatment was seen. In summary, we found no evidence of differences in the rates of diarrhea, aGVHD, graft failure, relapse and relapse-free survival, or OS at 1 year in GD-treated individuals vs. no-GD in our study.

**Table 2.** Exploratory comparison of clinical outcomes by randomized treatment arm.

|  | GD (N=10) |  | No GD (N=10) |  | p-value |
| --- | --- | --- | --- | --- | --- |
|  | N | % | N | % |  |
| Diarrhea in first 7 days post-HCT | 4 | 40 | 4 | 40 | 1.0 |
| Grade $\geq 2$ Acute GVHD | 1 | 10 | 3 | 30 | 0.58 |
| Grade 2 | 1 | 10 | 0 | 0 |  |
| Grade 3-4 | 0 | 0 | 3 | 30 |  |
| Bacteremia in first 100 days post-HCT | 1 | 10 | 5 | 50 | 0.14 |
| BSI episodes: | 1 |  | 8 |  | 0.09 |
| Gram positive episodes * | 1 |  | 5* |  |  |
| Gram negative episodes ^ | 0 |  | 3^ |  |  |
| Graft failure | 1 | 10 | 1 | 10 | 1.0 |
| Relapse of malignant disease # | 3 of 7 |  | 2 of 8 |  | ND |
| 1- year RFS: 73±11.4 (n=15 total) | | | | | |
| OS at 1 year | 10 | 100 | 10 | 100 | ND |

p-value based on Wilcoxon rank-sum test (Mann-Whitney *U* test).
Diarrhea defined as >3 stools per day.
\*One patient with 2 separate bloodstream infections (BSI) with MRSA 89 days apart, one patient with *Rothia dentocariosa* and *Enterococcus faecium*, one patient with *Staphylococcus epidermidis*.
^One patient with *E. coli*, one patient with *Leclercia adecarboxylata*, one patient with *Klebsiella oxytoca* (same patient who had two MRSA x2)

### Engraftment and immune reconstitution

The median time to neutrophil engraftment was 26 days (interquartile range (IQR), 23.5-29.2) in the GD arm and 24 days (IQR, 19.2-28.0) in the no-GD arm (p=0.47). One patient in the no GD arm had primary graft failure and underwent a second transplant on day +79. One patient in the GD arm had secondary graft failure and came off study at day +54. We also examined reconstitution of peripheral blood lymphocyte subsets in the two arms, excluding the two patients with graft failure. T-cell reconstitution was robust in both groups with median CD4+ T cell counts greater than 100 cells/μL at three months (median, 126.5 in GD arm vs 182.1 cells/μL in no-GD arm; p=0.24) and greater than 200 cells/μL at six months post-HCT (median, 206.5 in GD arm vs 248.2 cells/μL in no-GD arm; p=0.70) (Figure 3A). Within CD4+ T cells, the ratio of regulatory T cells to conventional T cells (Treg:Tcon) was similar between the study arms over the course of 12 months post-HCT (Figure 3B). Expansion of the naïve T cell fraction within CD4+ Tcon was also similar between the study arms (Figure 3D), suggesting that gut decontamination does not adversely affect thymic function. Recovery of CD8+ T cells and natural killer cells was similar between the study arms (Figures 3C and 3F, respectively). The absolute CD19+ B-cell concentration was significantly higher in the GD arm at 12 months with a median of 903.5 cells/μL (IQR, 814.0-984.8) compared to a median of 223.9 cells/μL (IQR, 190.1-333.3) in the no-GD arm (p=0.016) (Figure 3E). None of the patients in the no-GD arm received rituximab as part of their conditioning regimens or as part of post-transplant therapy. While it is difficult to draw conclusions based on this small and heterogeneous sample, the trends reported here can be investigated in a future larger study to further illuminate changes in immune reconstitution that are influenced by gut decontamination and the microbiota.

**Figure 3.**
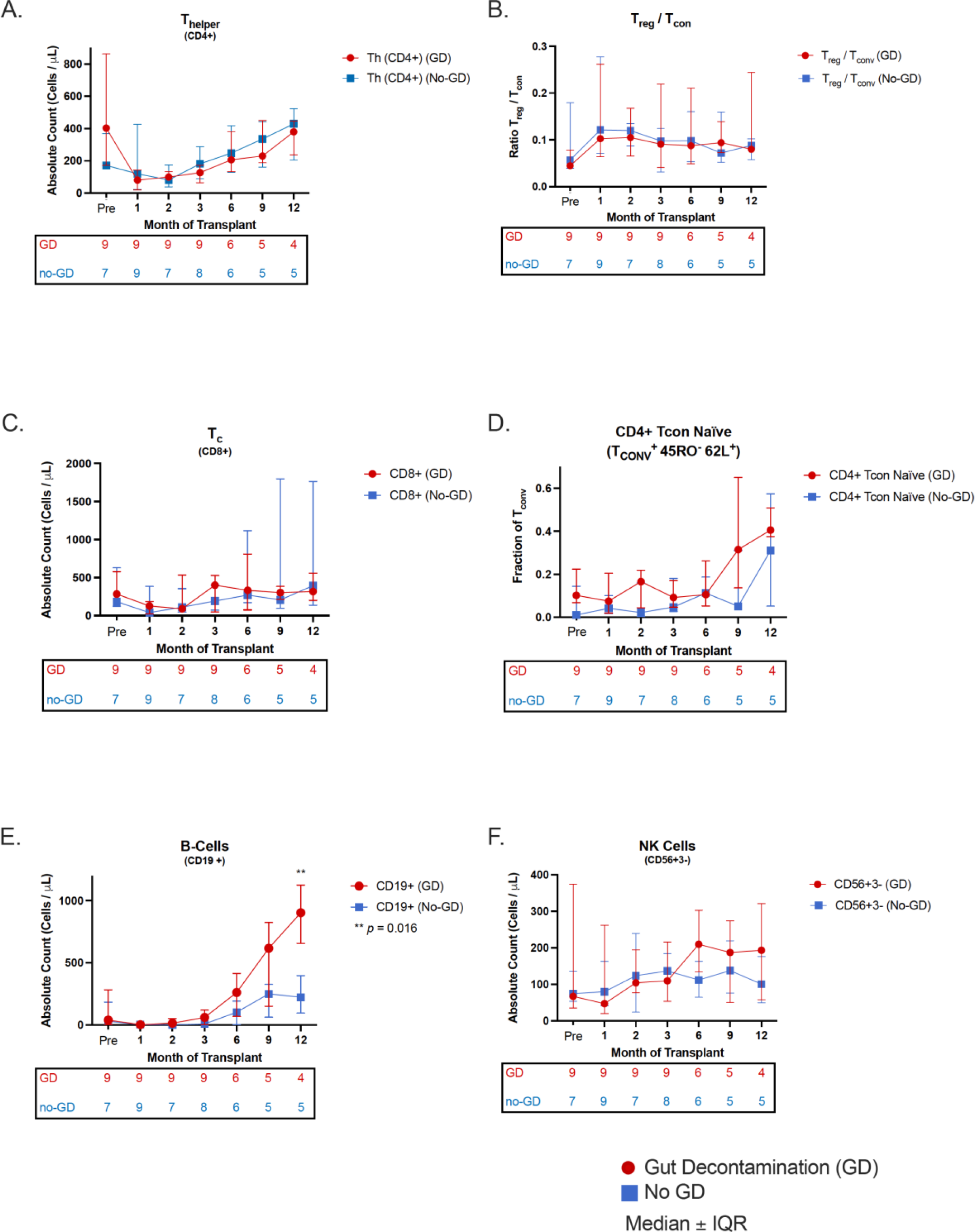
Immune reconstitution. Blood samples at pre-transplant, monthly for the first 3 months, then months 6, 9, 12. Shown are the median values ± interquartile range, along with the number of patients sampled at each time point below each graph. (A) CD4+ Th cells, (B) Treg:Tconv (C) CD8+ Tc cells, (D) CD4+ Tconv naïve cells, (E) CD19+ B-cells, ** *p*=0.016, (F) CD56+CD3- natural killer (NK) cells.

### Incidence of bloodstream infection (BSI)

In an exploratory analysis, we noted a trend of fewer BSIs in subjects enrolled in the GD compared to the no-GD arm. During the 100-day study period, a total of nine BSI episodes occurred in six patients, with eight episodes in the no-GD arm and one episode in the GD arm (*p*=0.09, Table 2 and Supplementary Table 2). The proportion of patients with BSI was 50% in the no-GD arm and 10% in the GD arm (Table 2, *p*=0.14). Five of the six patients had a BSI within the first 31 days. In a *post-hoc* exploratory comparison, the cumulative incidence of BSI was statistically significantly higher in the no-GD arm compared to GD (Figure 4, *p*=0.0483, Gray’s test). Seven of nine (78% of total) BSI episodes were from the no-GD arm and before the day of neutrophil engraftment.

**Figure 4.**
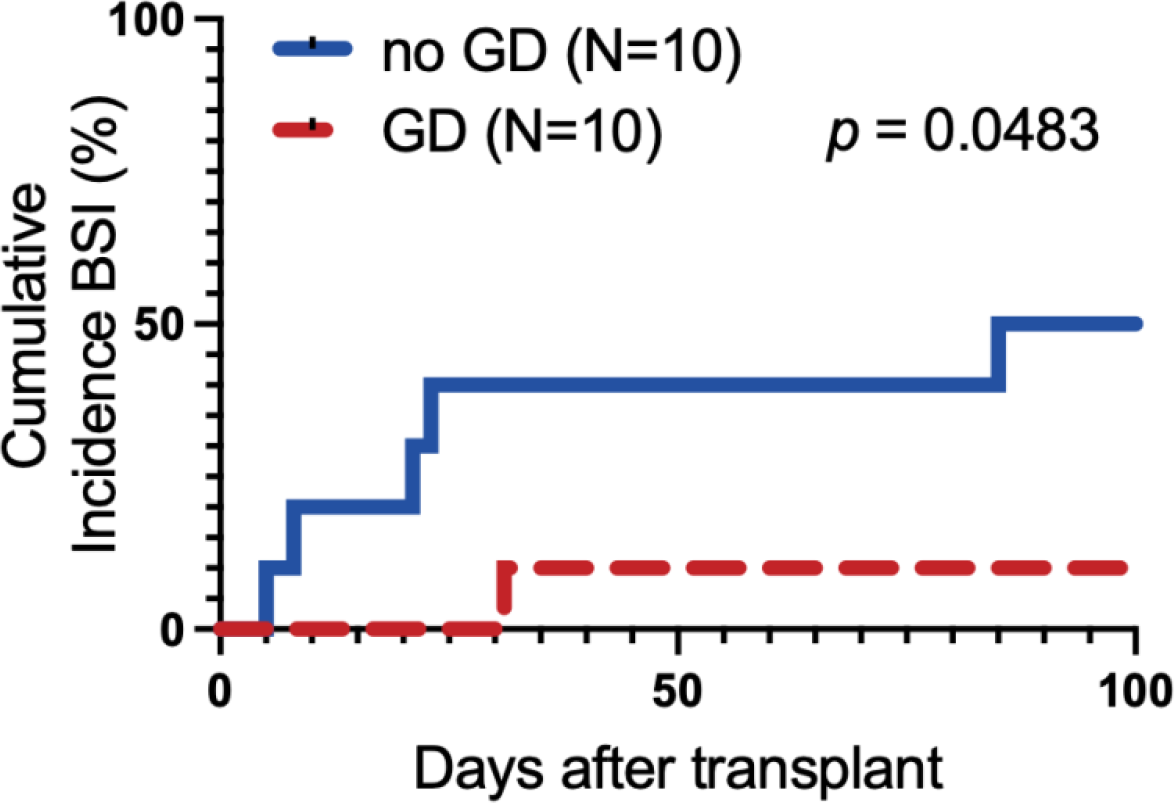
Cumulative incidence of BSI during the first 100 days of transplant. Patients are separated by treatment group with GD (dashed red line, N=10 subjects) and no-GD (solid blue line, N=10 subjects). Of the 6 patients with a BSI, 5 occurred within the first 31 days; one patient in the no-GD arm had BSI on day +85 relative to their first transplant (on day +6 of their second transplant). *p*=0.0483, Gray’s test.

### Species-level evidence that the BSI-causing bacterium is present in the gut microbiome

Given the interesting trend of increased BSI incidence in the no-GD vs. GD arm, we hypothesized that GD may decrease the burden of pathogens in the gut microbiota that can translocate across the mucosal barrier and subsequently cause a BSI. It is well established that the gut microbiome can be a reservoir of BSI-causing pathogens in this patient population [17, 20, 27, 36]; thus, we sought to test the hypothesis that patients in the no-GD arm would have BSI-causing pathogens within their gut microbiome before or during the time of infection.

In seven of nine of the BSI episodes, we were able to identify the species responsible for the BSI in the gut at a relative abundance of >0.1% within four days of the BSI (Figure 5 and Supplemental Figure 10, patients C03, C04 (two of three BSIs), C10, C11, C20, C22 (one of two BSIs)). We observed typically enteric bacteria causing a BSI in three patients (*Klebsiella oxytoca*, *Escherichia coli*, and *Enterococcus faecium* in patients C04, C10, and C22, respectively). Notably, three independent BSI episodes were caused by organisms in the genus *Staphylococcus*, which is typically categorized as non-enteric, non-mucosal barrier injury (non-MBI) related [37]. Interestingly, in each of these cases, *Staphylococcus* was found in the intestinal microbiota within four days of the BSI episode (Figure 5A and C, patient C04 (two independent BSIs 89 days apart, at 35% and 0.1% relative abundance) and C20 at 61% relative abundance). As a comparison, we see *Staphylococcus aureus* in <0.005% of the total reads with three independent healthy control stool samples (Supplemental Figure 5). In addition, we observed an increasing relative abundance of *Leclercia adecarboxylata* in the gut prior to the BSI in patient C03 (Supplemental Figure 10A).

**Figure 5.**
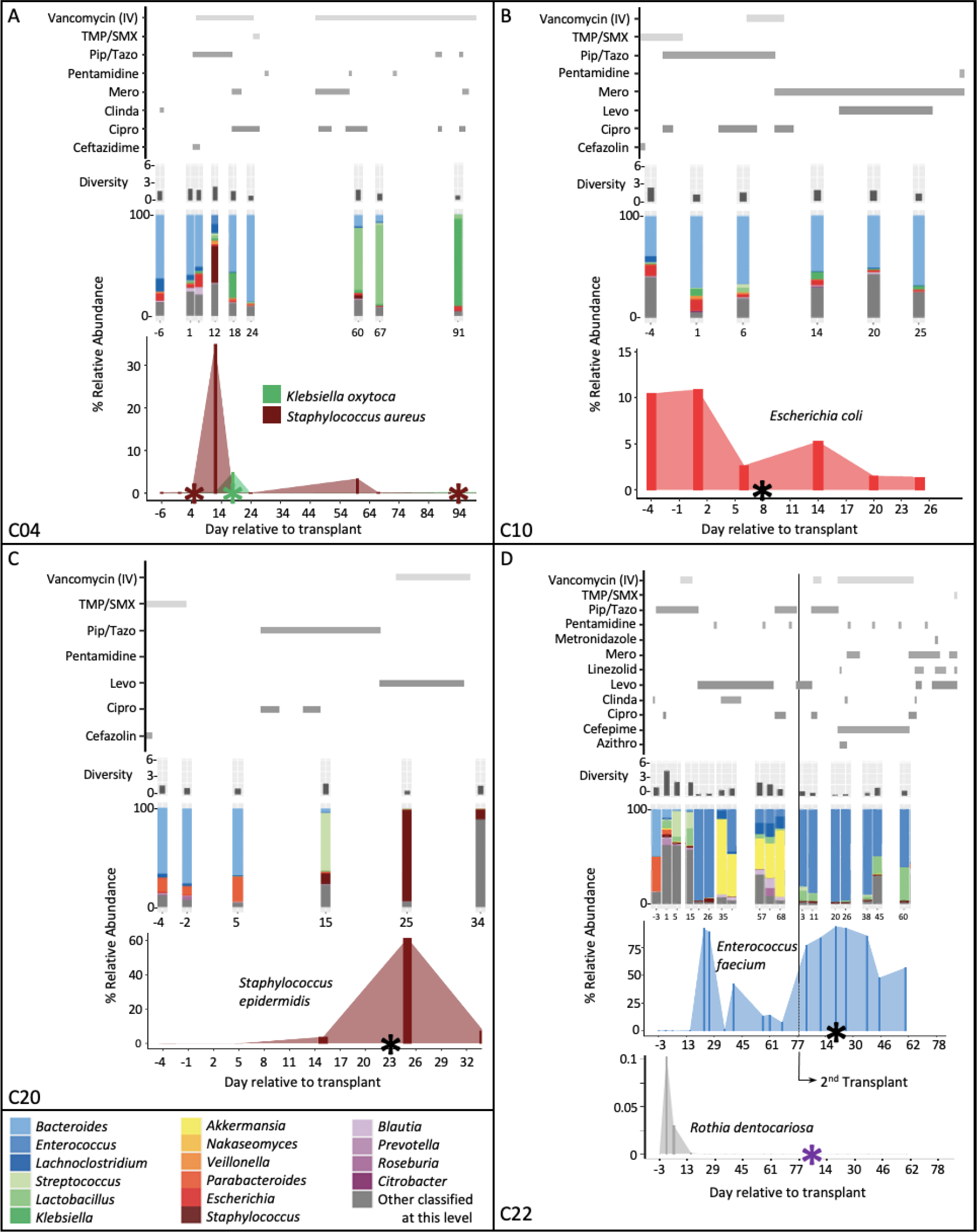
Bacterial abundance in the gut around the time of BSI. Multiple pathogens are present in the gut around the time of the BSI. Days relative to the course of the transplant on shown on the X-axis, (from top to bottom on the Y-axis) antibiotic administration, alpha (Shannon) diversity, relative abundance of microbes in the stool samples at the genus taxonomic level (with organisms listed by color according to the key at the lower left), and relative abundance in the gut of the BSI-causing organism with the date of the BSI shown as an asterisk (*). Note: Y-axis is a different scale between abundance plots for focused organisms found in the BSI. **(D)** Patient C22 received two transplants and had low abundance of *Rothia dentrocariosa*in the gut during the first transplant; *Rothia* was not detectable in the gut after day +15 of the first transplant. Antifungal and antiviral medications shown in supplemental figure 12. Information on patients C03 and C11 may be found in Supplemental Figure 10. Abbreviations: Azithro = Azithromycin, Cipro = Ciprofloxacin, Clinda = Clindamycin, Levo = Levofloxacin, Mero = Meropenem, PipTazo = Piperacillin / Tazobactam, TMP/SMX = Trimethoprim / Sulfamethoxazole (cotrimoxazole)

Two of the nine BSI episodes (a *Bacillus* bacterium and *Rothia dentocariosa*) are less likely to be derived from the lower gut (Supplemental Figure 10B and Figure 5D). While there was a rise in the relative abundance of the genus *Bacillus* in the stool sample (Supplemental Figure 10B, patient C11), the BSI isolate from C11 was subsequently sequenced and found to be *Lysinibacillus fusiformis* (also called *Bacillus fusiformis*, Supplementary Table 2), which was not detected in any of the stool samples of this patient. This suggests that either the BSI did not originate from the distal gut or was in sufficiently low quantity to preclude taxonomic identification at the species level. *Rothia dentocariosa*, an organism found predominantly in the oral cavity, was detected at low abundance in the fecal sample early in the first transplant of patient C22, but was undetectable after day +15, suggesting this BSI either originated from a location other than the distal gut, or was in low abundance in the lower gastrointestinal tract at the time of the BSI (Figure 5D, C22).

### Strain-level evidence that multiple BSI isolates are identical or nearly identical to those found in the gut microbiota around the time of the BSI

In the previous analysis, we found temporal concordance between species in the gut microbiota and the subsequent BSI, which suggested possible bacterial translocation across a damaged intestinal epithelium. If the BSI and gut strain were identical, this would be evidence of the gut being a likely source of infection. Thus, we sought to carry out a higher-resolution comparison to investigate for the presence of concordant strains of the BSI causing pathogens in the gut of patients. Because multiple strains of a given species can coexist in a gut microbiome, it is important to be able to assess multiple closely-related strains in a community and compare those to a single BSI strain. To carry out this type of strain analysis, we used *inStrain* [31], which can account for multiple strain populations in metagenomic sequencing data. Specifically, the population average nucleotide identity (popANI) metric only calls single nucleotide polymorphisms (SNPs) in positions where two samples do not share any alleles [31]. For example, two organisms of the same bacterial species will share >95% ANI, which is equivalent to five mismatches for every hundred bases [31]; two microbes are considered nearly identical if they have >99.9999% ANI, meaning there will one mismatch for every one million bases compared.

For this analysis, we generated whole genome sequencing data from 11 different BSI isolates from nine BSI episodes (Supplementary Tables 2 and 6); all strains analyzed are from no-GD patients except for one strain from patient C11 from the GD-arm. A median of 32.4x10^6^ (range 26.4 to 46.6x10^6^) raw read pairs were generated for each BSI isolate. After pre-processing (described in the methods), 15.6 x 10^6^ (range 7.5 to 23.5x10^6^) reads were obtained for each BSI isolate (estimated median 545-fold genome coverage). Draft genomes from BSI organisms were assembled and used as references for the *inStrain* comparison (assembly statistics in Supplementary Table 7). Sequencing reads from all stool and BSI isolates were mapped against a patient’s BSI draft genome, and samples with at least 20,000 mapping reads were retained. SNPs were called with *inStrain profile,* and pairs of samples were compared with *inStrain compare*. Pairs of samples that had at least 50% of the genome covered at a depth of at least five reads were considered for further analysis.

Going through the most notable cases individually (Table 3), no-GD patient C04 had two independent *Staphylococcus aureus* BSIs on day +5 and day +94 (Figure 5A). The *S. aureus* (at 35% relative abundance) in the stool sample from day +12 was identical to the BSI on day +5 (100% popANI, zero SNPs detected in 2.8 Mb of sequence compared (the size of the *S. aureus* genome is 2.8 Mb); Table 3, Supplemental Figure 11A). The *S. aureus* BSI on day +94 was nearly identical to the stool sample on day +12 (99.9999% popANI) with 2 SNPs between the samples. Patient C04 also had detectable *Klebsiella oxytoca* in the stool (0.3% relative abundance on day +12; 5% on day +18) and had a BSI with the same strain of *K. oxytoca* (100% popANI with 1.6Mb compared (the *K. oxytoca* genome is 6.02 Mb)) on day +18 (Figure 5A, C04). However, as *Klebsiella* was in lower abundance and not sequenced to high enough depth in samples prior to day +18, we were unable to determine whether the strain was present prior to BSI. Interestingly, while the clinical microbiology lab identified that the two *Klebsiella* strains from the blood culture had different sensitivity to ceftriaxone (Supplementary Table 6), the two isolates were identical by *inStrain* (100% popANI, zero SNPs) at the genomic level. This could be explained if there was a mobile genetic element or plasmid responsible for a difference in resistance.

**Table 3.**
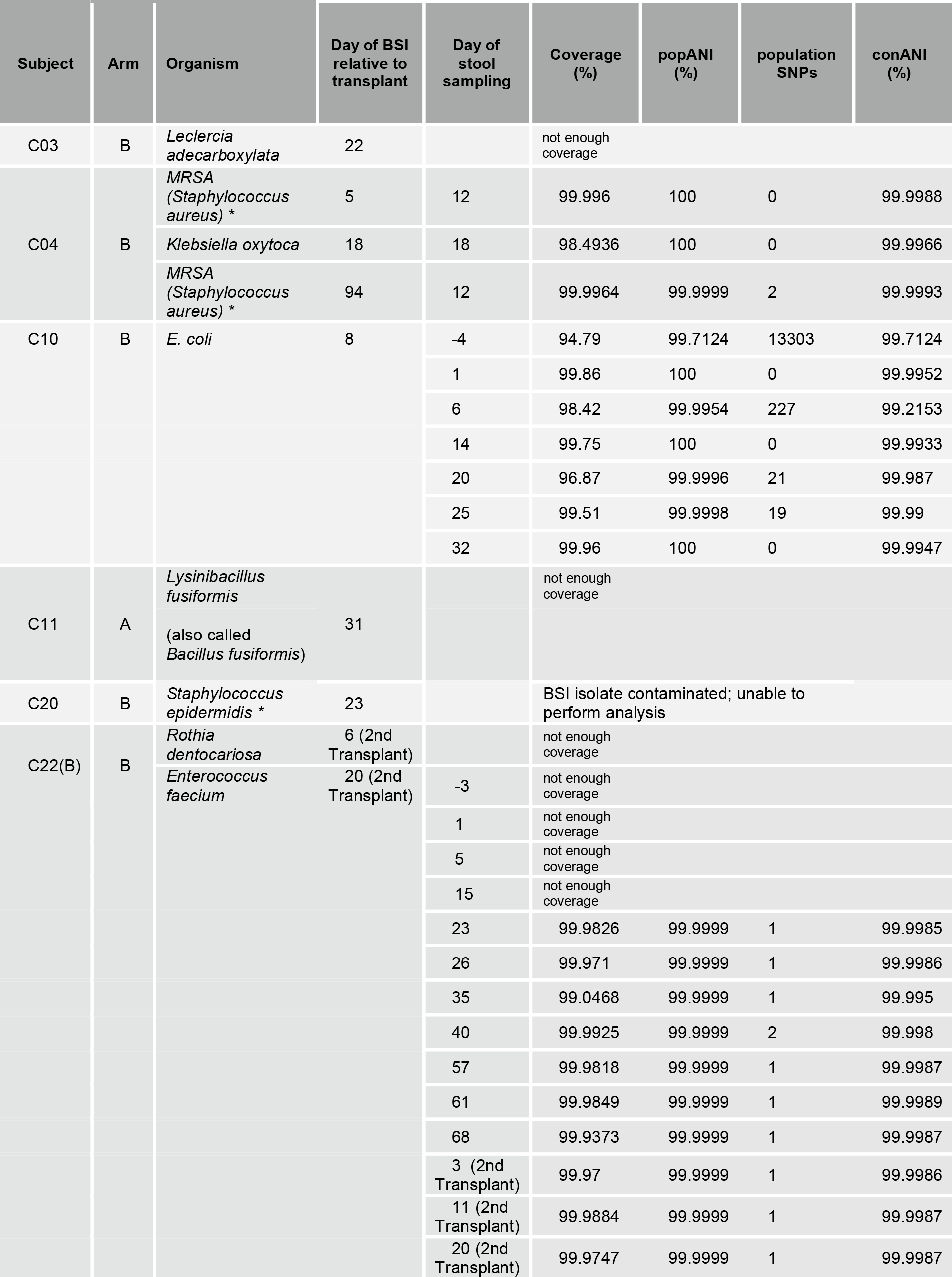
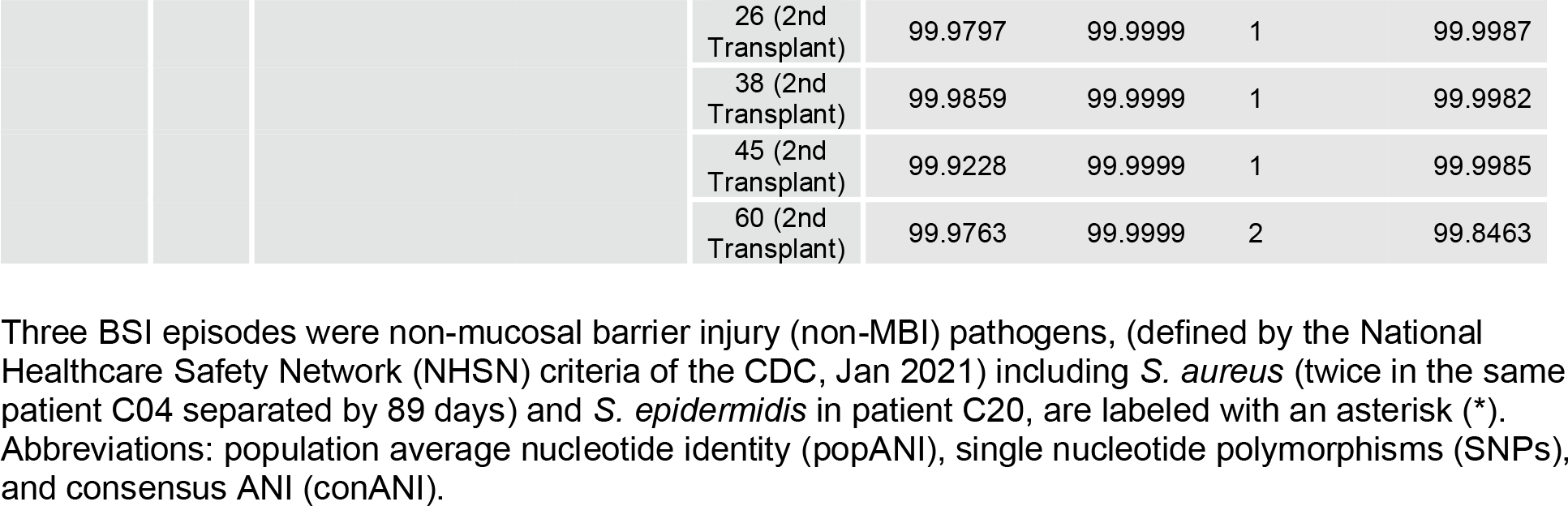
InStrain results comparing assembled BSI isolates to gut metagenomic samples.

Patient C10 in the no-GD arm had an *E. coli* BSI on day +8 (Figure 5B). An identical (100% popANI, 5 Mb compared (the *E. coli* genome is 5.12 Mb)) strain was found in the six stool samples collected from days +1 to +32. However, a different *E. coli* strain was present in the stool at day - 4 (99.7% popANI to BSI and other stool samples, with approximately 13,303 different SNPs, Table 3). Thus, at least two different strains of *E. coli* were observed in this patient at the different timepoints. It is possible that the BSI causing strain was present at day -4, but was simply below our limit of detection in this earlier sample. The two *E. coli* BSI samples on the same day (+8) from the clinical microbiology laboratory (Supplemental Table 6) were nearly identical to each other, with one SNP detected (99.9999% popANI).

Patient C22 in the no-GD arm experienced two BSIs after a second transplant (Figure 5D: *Rothia dentocariosa* on day +6 and *Enterococcus faecium* on day +20, relative to second transplant). In the eighteen stool samples from this patient, *R. dentocariosa* achieved a maximum of 0.1% relative abundance without enough sequencing coverage to conduct an *inStrain* comparison. By contrast, *E. faecium* in patient C22 was at >92% relative abundance at the time of the *E. faecium* BSI (Figure 5D, C22). In the samples from day +23 of the first transplant through the end of the study (14 samples total), *E. faecium* in the gut was nearly identical (>99.9999% popANI) to the BSI strain, suggesting that the same strain was present in this patient’s microbiota through the course of two transplants, and likely eventually caused BSI (Figure 5D, Table 3).

In patients C03 (no-GD arm) and C11 (GD arm), the organism (*Leclercia adecarboxylata and Lysinibacillus fusiformis*, in C03 and C11 respectively) found in the BSI was either undetectable or at low abundance and did not have sufficient sequencing depth and coverage in the stool samples to make a conclusion regarding strain specificity using the *inStrain* comparison (Table 3). No-GD patient C20 experienced a *Staphylococcus epidermidis* BSI on day +23, with a rise in relative abundance in the gut microbiota from 3.7% on day +15 to 61% on day +25 (Figure 5C, C20), suggesting the BSI originated from the gut, however, we were unable to do strain-level analysis as the original BSI-causing isolate was archived, but upon sequencing was identified to be *E. coli*, a likely contaminant of the culture (Table 3).

Based on these findings, at least five of the nine BSIs in our study are identical or nearly identical to a species found in the gut microbiota using the popANI genome comparison of *inStrain*. Two additional BSIs (*Leclercia adecarboxylata* from patient C03, and *Staphylococcus epidermidis* from patient C20) may also have originated from the gut based on an increase in the relative abundance of the bacteria around the time of the BSI, although strain level confirmation of this prediction is lacking. This suggests that in up to seven of the nine BSIs, the infection-causing pathogen is present in the gut in patients from the no-GD arm; by contrast none of the BSI-causing pathogens are present in the gut in patients from the GD arm (Figure 5, Supplemental Figures 10 and 11). Collectively, these data demonstrate that the gut microbiota is a reservoir for pathogens traditionally derived from the gut, and that microbes like *Staphylococcus*, that are not typically considered as normal gut bacteria, may subsequently cause a gut-derived BSI in pediatric allo-HCT patients.

## DISCUSSION

Due to the lack of evidence supporting a clear benefit, GD is not recommended as a standard practice for the prevention of aGVHD or bacteremia. An informal survey of transplant centers in the United States in 2017 indicated that approximately 40% of adult and pediatric centers routinely practiced GD [7]. As part of this study, we surveyed 101 pediatric HCT centers in the U.S. and Canada regarding their GD practices in 2019. Of the 32 centers that responded, only a small proportion (3 of 32; 9.4%) of centers were using GD as part of the aGVHD prophylaxis regimen (Supplemental Document 1). The practice of GD in patients undergoing allo-HCT is based on murine data, as well as early single-arm and retrospective studies suggest that using GD to alter the intestinal microbiota may protect against aGVHD [2, 5, 8]. In this study, the first prospective randomized trial of GD in pediatric allo-HCT recipients, we directly investigate how GD with oral vancomycin-polymyxin B alters the microbial composition on a species and strain level using NGS. We also characterize secondary clinical outcomes and identify BSI-causing pathogens traceable to the gut either temporally or by strain-specific analysis (concomitant gut colonization) via strain-level comparative genomics.

We show that there is no appreciable difference in Shannon diversity of the digestive tract during the peri-transplant period between GD and no-GD arms. While subjects in the GD arm had variable administration of vancomycin-polymyxin B (supplemental fig. 2), the changes in diversity at 2 weeks did not correspond to the amount of GD the individual subject received. Furthermore, GD does not appear to lead to a decreased use of systemic antibiotics, as there was not a statistically significant difference in the time of exposure to prophylactic and therapeutic antibiotics between the two arms to account for the similarity in Shannon diversity. While Shannon diversity is sensitive to loss of rare taxa [34], a larger sample size with an alternative measure of changes in individual taxa over time may account for any differences between the GD and no-GD arms.

For example, we considered alternative analysis including the differential abundance of species over time using a mixed effect linear model, however our analysis is limited by the heterogeneity and large numbers of taxa relative to the small number of samples.

Despite promising evidence from earlier studies showing that non-absorbable antibiotics were associated with a lower incidence of GVHD [1, 5], more recent data suggests a more complex picture. For example, one retrospective report suggests that choice of antibiotics is critical, as cefuroxime, tobramycin, and nystatin in the GD arm was associated with an increased risk of developing aGVHD compared to non-GD [38]. Several studies have expanded this concept showing that systemic broad-spectrum antibiotics and loss of microbial diversity (specifically commensal organisms) are associated with gastrointestinal (GI)-GVHD and GVHD-related mortality [10, 12]. While our study is not powered to assess a difference in aGVHD incidence, we found that three patients had grade III-IV aGVHD in the no-GD arm vs one patient in the GD with grade II aGVHD. We also note that the GD arm had more matched related donors compared to the no-GD arm, which could explain any trends observed. Collectively these data leave an open question as to whether GD decreases the risk of grade III-IV aGVHD, which will need to be assessed in a larger prospective trial.

Based on prior studies suggesting interactions between the gut microbiota and development of circulating immune cells, we examined immune reconstitution of T- and B-cell subsets over the first year post-HCT. Although the reconstitution of T-cell subsets was similar between the study arms, the absolute CD19+ B-cell count at 12 months was significantly higher in the GD arm. This difference cannot be explained by *in vivo* depletion as none of the patients in the no-GD arm received rituximab post-HCT. Several reports demonstrate extensive crosstalk between the microbiota and B-cell diversity [39–41], and that early B-lineage development in mice is influenced by the gut microbiome [42]. While difficult to draw inferences with the small numbers here, an analysis of immune reconstitution and potential implications for the risk of infection will need to be fully characterized in a larger study.

While the impact of GD on infection-related outcomes was not part of our pre-specified primary or secondary analyses, we postulated that GD with oral vancomycin-polymyxin B may have an impact on decreasing the rate of BSIs that originated from the gut compared to no-GD (Supplemental Figure 13). In an exploratory analysis, we found that all BSIs with concomitant localization to the gut were observed in the no-GD arm. Interestingly, historic publications reporting on GD demonstrated that many of the Gram-negative bacteria isolated from the BSIs were still sensitive to the drugs used for decontamination [22], and meta-analysis in the ICU demonstrated that there was no increase in antimicrobial resistance with selective decontamination [43]. The primary mechanism of GD reducing bacteremia in critical care patients is thought to occur by limiting the growth of select bacteria, including opportunistic bacteria such as *Enterococcus* and from the phylum proteobacteria [44], which includes the genera *Escherichia* and *Stenotrophomonas*. No prospective study in pediatrics has been conducted to date with a standard of care arm that is appropriately powered to address if GD decreases the risk of BSI. Furthermore, there have been very limited studies to compare systemic antibiotic prophylaxis to non-absorbable gut decontamination (including Gluckman, E., *et al.,* [45], and reviewed in a meta-analysis by Kimura, *et al.* [46]). The largest study in pediatrics to show a reduction in bacteremia was conducted in children undergoing initial therapy for acute leukemia and used levofloxacin for systemic prophylaxis, rather than gut decontamination; a second arm of the study did not reach the level of statistical significance for children undergoing HCT (Children’s Oncology Group Study ACCL0934) [47]. Given the above findings, there is insufficient data supporting a clear benefit of GD in humans in HCT at this time. However, a larger multi- center trial may inform the question of whether oral vancomycin and polymyxin B decreases the burden of pathogens in the gut microbiota that can then translocate across the mucosal barrier and subsequently cause a BSI.

HCT patients often have injured mucosae secondary to conditioning chemotherapy and neutropenia, increasing their risk of mucosal barrier injury-laboratory confirmed bloodstream infection (MBI-LCBI). Due to their immunocompromised status and central venous catheters, HCT patients are also at risk for non-MBI-LCBI infections, including central line-associated BSI (CLABSIs). This is an important distinction as MBI-LCBI’s are not prevented by improved central venous catheter care when compared to CLABSI [13, 48–50]. Given that there is approximately 18% mortality attributable to BSI (range 12% to 20%) in this patient population [13], being able to appropriately identify and subsequently develop methods to decrease this rate could have a profound impact on patient survival during HCT. We demonstrate here that the bacterial species responsible for the BSI are found in the gut in seven of nine BSI episodes. While many of those identified in the gut microbiome are Gram-negative enteric organisms, we also found evidence of classically non-MBI-LCBI organisms such as *Staphylococcus* species in the gut microbiome. These data, along with previous work by Tamburini, *et al.* [27], Kelly, *et al.* [20], and Zhai, *et al.* [36], strongly suggest that the gut can be a reservoir for pathogens in HCT recipients of enteric origin and that have traditionally been classified as non-MBI pathogens, such as *Staphylococcus aureus and epidermidis*. Overall, there may be skin- and nares-resident organisms such as *S. aureus* and *S. epidermidis* that can colonize and grow to high relative abundance in the gut microbiomes of severely immunocompromised hosts. Collectively, these data provide a basis to inform future studies about potential methods to decrease BSIs in allo-HCT patients.

While the findings presented here suggest that GD is generally safe and has a relatively limited impact on the gut microbiome, there are several limitations to our study. First, the results are from a single center study and the sample size is relatively small. While originally powered to detect a difference of gut microbiome Shannon diversity of 4.0 in the no-GD arm and 2.8 in the GD arm, the magnitude of the change was less than anticipated. Some of this disparity is likely due to confounding variables like systemic antibiotics creating an even larger heterogeneity of the microbiota data than originally anticipated. Further adding to the interpretation of the data is the variability in administration of the oral vancomycin and polymyxin B, which is often challenging to administer due to palatability of the antibiotics. This may be partially alleviated by the practice of nasogastric tube insertion for feeding and administration of oral medications early in the transplantation course (e.g., around the time of transplant and several days before the onset of mucositis). Nevertheless, the ability to take the GD antibiotics may complicate future efforts to carry out a prospective trial and limits our ability to conclude that GD had no effect on the primary and secondary outcomes. We also do not consider the microbiota from other sites in the body (e.g., skin) which patients may be extensively colonized with and thus cannot say definitively that the skin or other sites were not the source of the BSI.

Despite these limitations, the prospective design of this study, the standardized collection of samples, the application of NGS technologies and the use of *inStrain* to compare BSI genomes and fecal metagenome assemblies (MAGs) enable us to further investigate the consequences of GD vs. no-GD. In particular, whole genome sequencing of stool and bacterial isolates, along with use of the *inStrain* algorithm, allows for the confident and accurate comparison of bacterial isolates and metagenomes to identify identical or nearly identical microbial strains. In addition, by using a genome-wide comparison, we provide the framework to subsequently interrogate strain specificity which can have important impacts on antibiotic resistance and pathogenicity of particular organisms (e.g., exploiting metabolic pathways of specific strains). A larger study could test the model that GD decreases the number of, or eliminates entirely, bacteria that can potentially be pathogenic (Supplemental Figure 13) and translocate across the mucosal barrier.

Ultimately, we aim to identify specific patterns and determine ways that we can alter the gut microbiome to decrease the risk of bloodstream infections that originate from the gastrointestinal tract. For example, efforts have been made to use the microbiota composition to predict the risk of a BSI [19, 21] and multi-drug resistant BSI [51]. Whether identification and subsequent targeting of microbes can be achieved to decrease the risk for BSI in the allo-HCT population remains an open question. Identification of patients who are at an increased risk of BSIs from the gut could lay the groundwork to develop microbiota-based therapies to decrease the abundance and/or prevalence of pathogens in the gut microbiota, including appropriate antibiotic stewardship, systemic prophylaxis, gut decontamination, phage therapy, fecal microbiota transplantation (FMT) [52], and other microbial- or host-modifying treatments including prebiotics [53]. In conclusion, in this phase 2 RCT of 20 patients, we noted that all BSIs traced to the gut were found in the non-GD arm. Furthermore, *Staphylococcus* was found in the gut suggesting that expanding the number of organisms defined as MBI-LCBI for allo-HCT patients is an important step for detecting and subsequently designing ways to mitigate the risk of BSI during HCT. Finally, these data suggest that oral vancomycin-polymyxin B GD may protect against post-HCT BSI by decreasing the prevalence or abundance of pathogens that can translocate across the mucosal barrier and subsequently cause gut-derived BSIs, a finding that will need to be verified in a larger trial.

## METHODS

### Cohort selection and study design

We performed a randomized phase 2 trial (+/- gut decontamination, ClinicalTrials.gov Identifier: NCT02641236, CONSORT flow diagram in Supplemental Figure 1) examining the impact of gut decontamination (GD) with oral vancomycin-polymyxin B on intestinal microbiota composition of allogeneic HCT patients compared to no-GD. Eligibility included any recipient, ages ≥ 4 years to 30 years (Adult: 18-30 years, pediatric: 4-17 years) and toilet-trained, of 9/10 or 10/10 (HLA-A, - B, -C, -DRB1, -DQB1) matched bone marrow allogeneic-HCT, or 4/6, 5/6 and 6/6 (HLA-A, -B, - DR) matched cord blood allogeneic HCT. Stool from a cohort of two healthy sibling donors were collected as a comparison group (ages ≥ 4 years and toilet-trained). Detailed eligibility criteria are available in Supplementary Document 2, “Clinical inclusion and exclusion criteria.” Patients were enrolled and were randomized (1-to-1) to either “GD” arm or “no-GD” arm. The primary endpoint was microbial (Shannon) diversity at two weeks post-HSCT. The trial was powered to detect a difference in Shannon diversity index of 1.2 (4.0 for no-GD vs 2.8 for GD) with a one-sided t-test and alpha=0.05, and assuming a standard deviation of 0.9. An intention-to-treat comparison of GD vs no-GD was performed using the Wilcoxon rank-sum test. Participants assigned to the GD arm received non-absorbable, oral vancomycin-polymyxin B capsules according to body surface area; 375mg/m2 BSA of vancomycin and 187mg/m2 of polymyxin B (see Supplemental Table 1 for details). Random allocation of anonymous identifier, enrollment, and assignment completed under the supervision of the principal investigator (J.S.W.) of the clinical trial. Oral gut decontamination began on day -5 relative to the hematopoietic cell infusion date (day 0) and continued through neutrophil engraftment, defined as an absolute neutrophil count ≥ 500 cells/mm^3^ for three consecutive days. Patients in no-GD received the institutional standard of care including all other supportive care as did patients in the GD arm. Antifungal and antiviral prophylaxis was used at the discretion of the treating physician generally starting at day -9 (e.g., fluconazole) and day -5 (e.g., acyclovir) respectively (administration data in Supplemental Figure 12). Use of any agent (e.g. sulfamethoxazole/trimethoprim, pentamidine) for prophylaxis of *Pneumocystis jirovecii* pneumonia was permitted. Secondary endpoints include the frequency of diarrhea in the first seven days post-HCT, incidence of grade ≥ 2 acute graft versus host disease (aGVHD) during the first 100 days post-transplant, survival, malignant disease relapse at one year after study entry, progression free survival (defined in this study as time from randomization to the earlier of progression of malignant disease or death due to any cause at one year after study entry), and immune reconstitution. Exploratory outcomes include bacteremia during the first 100 days post-transplant. Clinical records including details of the transplant type, conditioning regimen and prophylaxis medications for aGVHD, antibiotic administration, microbiological information (including BSI and antibiotic resistance data), clinical symptoms (including aGVHD, diarrhea defined as >3 stools per day), and outcomes (relapse, death, and aGVHD) were obtained from the patient chart.

Stools samples were collected and stored immediately at 4°C and frozen at -80°C in cryovials within 24 hours of collection. Stool samples were collected as follows: weekly (+/- 3 days) prior to neutrophil engraftment; monthly (+/- 2 weeks) after neutrophil engraftment until six months; at six months and one year (+/- 1 month); within 48 hours of aGVHD diagnosis or positive blood culture.

### Flow cytometry and immune profiling

Phenotypic analyses of lymphocytes subsets were performed at: pre-transplant, 1, 2, 3, 6, 9, and 12 months after transplant. Briefly, 50 µL of EDTA whole blood was subjected to red blood cell lysis using 1x BD PharmLyse (BD Biosciences) and subsequently incubated with fluorochrome-conjugated monoclonal antibodies with individual subsets enumerated in a FACSCanto II flow cytometer and analyzed using BD FACSDiva (both from BD Biosciences) and FlowJo software (TreeStar) as described previously [54]. CD4Treg were defined as CD3+CD4+CD25^med- high^CD127^low^; CD4Tcon as CD3+CD4+CD25^neg-low^CD127^med-high^; B cells as CD19+, naïve B-cells as CD19+CD27^neg^, memory B-cells as CD19+CD27+; and natural killer (NK) cells as CD56+CD3-. Within CD4Treg and CD4Tcon, subsets were defined as follows: naïve T cells (CD45RO- CD62L+), CM (CD45RO+CD62L+), and EM (CD45RO+CD62L-).

### BSI antibiotic susceptibility testing

Antibiotic susceptibility testing on isolates from bloodstream infections was performed by the Clinical Microbiology Laboratory at Boston Children’s Hospital (BCH) / Dana-Farber Cancer Institute (DFCI), except for colistin (polymyxin E), which was performed at Stanford Health Care Clinical Microbiology Laboratory using the disk elution test as previously described [55]. Minimal inhibitory concentrations (MIC) for Enterobacteriales were interpreted using breakpoints according to Clinical and Laboratory Standards Institute (CLSI) [56] and colistin (polymyxin E) using the European Committee on Antimicrobial Susceptibility Testing (EUCAST) [57].

### BSI isolate cultures

BSI isolates from HCT patients who received care at BCH and DFCI were obtained from the Clinical Microbiology Laboratory at DFCI. Isolates were subsequently grown on agar slants, transferred to Luria-Bertani (LB) broth, and grown to saturation at 37°C. Bacteria were pelleted by hard spin (10,000 g) followed by removal of the supernatant and frozen at -80°C until DNA extraction.

### DNA extraction and whole genome shotgun (WGS) metagenomic sequencing of stool samples and BSI isolates

Genomic DNA was extracted from patient stool samples and BSI cultures using the QIAamp Fast DNA Stool Mini Kit (QIAGEN, Cat. #19593) per the manufacturer’s instructions with the following modifications: in suspension buffer, samples were heated to 95°C and subjected to seven rounds of bead-beating for 30 seconds, alternating with cooling on ice for 30 seconds prior to addition of proteinase K and lysis buffer. DNA concentration was measured using Qubit Fluorometric Quantitation (DS DNA High-Sensitivity Kit, Cat. #Q32851, Thermo Fisher Scientific). DNA sequencing libraries were prepared using the Nextera XT DNA Library Prep Kit (Illumina) and Nextera FLEX (Illumina) for samples unable to be prepared with Nextera XT due to low biomass. DNA library fragment length distributions were quantified via Bioanlyzer 2100 instrument (Agilent Technologies) using the High Sensitivity DNA kit (Cat.#5067-4626, Agilent Technologies) per the manufacturer protocol.

Libraries were pooled with unique dual sample indices to avoid barcode swapping [58] and sequenced on Illumina HiSeq4000 or NovaSeq P150 platforms with a read length of 150bp. Sequencing performed by Novogene (Sacramento, CA).

For microbiota analysis, a healthy donor stool was used as a positive control for batch -to-batch variation, a known microbiota community (ZymoBIOMICS microbial community standard, Cat#D6300) to determine microbial extraction bias, and a negative control carried through the entire DNA extraction, library preparation, and sequencing.

### Computational methods

#### Preprocessing

Fecal and BSI short-read WGS metagenomic sequencing reads were preprocessed to remove sequencing adapters, PCR artifacts and duplicate reads, and any reads mapping to the human genome, using established workflows available at: https://github.com/bhattlab/bhattlab_workflows/blob/master/manual/preprocessing.md [59].

Briefly, sequenced reads were deduplicated using SuperDeduper [60] and trimmed using TrimGalore v0.6.5 [61] with a minimum quality score of 30 for trimming and minimum read length of 60. All reads that aligned to the human genome (hg19) were removed using BWA v0.7.17 [62] with final results of pre-processing read-counts shown in supplemental figure 4. Sequences then underwent quality control using FastQC v0.11.9 [63]. Bioinformatics workflows were implemented using Snakemake [64].

#### Classification with Kraken2 and diversity calculations

Short-read data was taxonomically classified using Kraken2 [32] against a database of all bacterial, fungal, and viral genomes in the NCBI GenBank database assembled to complete genome, chromosome, or scaffold quality as of January 2020. Species abundance was estimated using the Bracken [65] database, built using a read length of 150 and *k-mer* length of 35. Workflows are available at: https://github.com/bhattlab/kraken2_classification [66]. Diversity of the microbes in the samples was calculated using the R package Vegan v2.5-7 [67].

#### Assembly and binning

Short-read sequences from stool samples and BSI isolates were assembled using SPAdes v3.15.2 [68]. Stool metagenomic sequences were subsequently binned using CONCOCT v1.1.0 [69], Metabat2 v2.15 [70], and Maxbin v2.2.7 [71], aggregated using DASTool v1.1.1 [72] and de- replicated using dRep v2.5.4 [73]. Bins were evaluated for completeness and contamination using QUAST [74]. Metagenome assembled genome (MAG) quality was assessed using previously established standards by Bowers, *et al.* [75] and Nayfach, *et al.* [76]. Workflows are available at: https://github.com/bhattlab/bhattlab_workflows/blob/master/manual/assembly.md [59].

#### Antibiotic resistance gene detection

Assembled BSI contigs and binned contigs from stool metagenomic sequences were profiled for antibiotic resistance genes (ARGs) with the Comprehensive Antibiotic Resistance Database (CARD) and the Resistance Gene Identifier (RGI) using default parameters [77].

#### Determining strain specificity of BSI isolates and stool metagenome assemblies

To compare bacterial strains in multiple samples, including gut metagenomes which may have multiple strains of the same species present, we used inStrain v1.0.0 [31]. Sequencing reads from multiple samples were mapped against assembled BSI genomes using BWA [62]. Pairs of samples with >50% coverage breadth at a depth of at least five reads were compared to analyze SNPs and determine average nucleotide identity (ANI) between the samples.

### Data availability

All sequencing data sets from the current study have been deposited in the Sequence Read Archive under the National Center for Biotechnology Information (NCBI) BioProject ID PRJNA787952 at http://www.ncbi.nlm.nih.gov/bioproject/787952

### Statistics

#### Statistical analysis and graphical presentation methods

Taxonomic abundance plots, antibiotic time course, and vancomycin-polymyxin B dosage graphs were created using GraphPad Prism version 9.1.2 for macOS, GraphPad Software, San Diego, California USA, www.graphpad.com, and the ggplot2 package v3.3.3 [78] with code modified from [20] and [79]. Comparisons by treatment group were performed using Fisher’s exact test (for binary variables), Wilcoxon rank-sum test (for continuous variables), or Wilcoxon signed-rank test (for paired continuous variables) for comparison of baseline versus two-weeks post-HSCT within patients. The Wilcoxon rank-sum and Wilcoxon signed-rank tests were adjusted using a false-discovery rate (FDR) ≤0.05. Cumulative incidence curves of BSI were compared using the Gray’s test with adjustment for the competing risk of death. Alpha and beta diversity were calculated using the vegan package v2.5-7 [67], and compared with the Wilcoxon rank-sum test and corrected using an FDR of ≤0.05. Analysis of Similarity (ANOSIM) statistic after 999 permutations was done for comparison of beta-diversity for patients with healthy sibling samples to compare. Plots were generated using R as previously reported [20, 27]. Relative abundance plots of specific taxonomic groups were generated using Animalcules R interface [80]. Figure 1 and graphical abstract created with BioRender.com

#### Study approval

The trial was approved by the institutional review board (IRB) under the IRB of Dana-Farber Cancer Institute (DFCI) (Protocol #15-394 approved Oct. 2015; principal investigator: J.S.W.), was performed at Boston Children’s Hospital (BCH) and DFCI. IRB protocol was open to patient entry March 2016 through September 2019. Informed consent for the patient (if ≥18 years), parent (if <18 years), or legally authorized representative, was obtained prior to any specimen collection. Full protocol available at DFCI. Trial is registered under ClinicalTrials.gov Identifier: NCT02641236.

## AUTHOR CONTRIBUTIONS

J.S.W., A.S.B., and J.R. conceived of the original study.

W.B.L and J.S.W. designed the clinical trial.

L.E.L., S.P.M., and C.N.D. contributed to the clinical protocol, assisted in enrollment and acquisition of clinical samples.

S.Silverstein, S.K., and O.B. collected the clinical samples.

Survey Core at DFCI https://surveycore.org/ conducted the informal survey of gut decontamination practice at pediatric HCT centers.

C.J.S., J.S.W., W.B.L., and N.C. organized, generated figures, and analyzed the clinical data. C.J.S., M.M.L., extracted DNA, and prepared short-read sequencing libraries, selected samples for sequencing.

C.J.S. B.A.S., and A.S.B. analyzed the sequencing quality.

C.J.S., B.A.S., and A.S.B. conceived of the assembly approach and performed the InStrain analysis.

N.B. and A.L. performed and analyzed antibiotic resistance of gram-negative BSI’s to polymyxin B / colistin.

C.J.S. and S.TJK. wrote and modified code and generated figures for the clinical data.

C.J.S. and T.M.A. analyzed the infectious disease data.

S.Sun and A.A.F., modeled and considered alternative analysis paths for the microbial sequence datasets.

C.J.S. carried out all analyses, wrote the manuscript, and generated figures & tables. All authors reviewed and commented on the manuscript.

The authors read and approved the final manuscript.

## Data Availability

All data produced in the present study are available upon reasonable request to the authors.
All sequencing data sets from the current study will be deposited in the Sequence Read Archive under the National Center for Biotechnology Information (NCBI) BioProject ID PRJNA787952 at http://www.ncbi.nlm.nih.gov/bioproject/787952 (data release will occur at time of publication)

## ACKNOWLEDGEMENTS

We thank the patients and their families, along with the hard work and dedication of the clinical and clinical-research teams, without whom this research would not be possible.

We appreciate the members of the Bhatt lab and K. Weinberg for their comments and suggestions on this work, including S. Zlitni, A. Han, A. Natarajan, P. West, Y. Pinto, R. Chanin, M. Chakraborty, and D. Maghini.

This work utilized computing resources provided by the Stanford Genetics Bioinformatics Service Center, supported by NIH S10 Instrumentation Grant S100D023452. Computing costs were supported via a NIH S10 Shared Instrumentation Grant 1S10OD02014101.

C.J.S. is a Pete and Arline Harman Fellow with the Stanford Maternal & Child Health Research Institute (MCHRI) and supported by the MCHRI and T32-DK098132.

A.S.B. is supported by Damon Runyon Clinical Investigator Award, V Foundation Scholar award, Sloan Foundation Fellowship, Emerson Collective grant, NIH R01 AI143757 and R01 AI148623.

The content is solely the responsibility of the authors and does not necessarily reflect the official views of the NIH.

Interim analyses have been previously reported in abstract form: *Blood* (2019) 134 (Supplement_1): 5665 https://doi.org/10.1182/blood-2019-122480

## Disclosures and potential conflict of interest

Except for those listed below, the authors have declared that no potential conflict of interest exist.

B. Siranosian:

Employee: Cellular Longevity Inc. Consultant: Imago Biosciences.

S. Margossian:

Consultant: Novartis Salary: CueBio Inc

J. Ritz:

Consultant: Akron Biotech, Avrobio, Clade Therapeutics, Garuda Therapeutis, LifeVault Bio, Novartis, Rheos Medicines, Talaris Therapeutics, TScan Therapeutics.

Research funding: Amgen, Equillium, Kite/Gilead, Novartis

A. Fodor:

Consultant: Gelesis, ByHeart and L-nutra

W. London:

Consultant: Jubilant Draximage and Merck

A. Bhatt:

Scientific Advisory board: ArcBio, Caribou Biosciences Consultant: Kaleido Biosciences, biomX

Research collaboration without funding support: 10x Genomics, Illumina, Oxford Nanopore Technologies

Nonprofit Board: Global Oncology, Inc.

J. Whangbo:

Employee: Vor Biopharma

**Trial Registration.** ClinicalTrials.gov NCT02641236

