## Supplemental Tables and Figures for "Randomized controlled trial of gut decontamination in pediatric patients undergoing allogeneic hematopoietic cell transplantation"

| | ORCID |
|---|---|
| Christopher J. Severyn | 0000-0002-0884-6372 |
| Benjamin A. Siranosian | 0000-0002-4935-8172 |
| Sandra Tian-Jiao Kong | 0000-0002-0044-562X |
| Angel Moreno | |
| Michelle M. Li | 0000-0003-0223-7485 |
| Nan Chen | |
| Shan Sun | 0000-0003-0349-2664 |
| Tessa M. Andermann | 0000-0002-0844-7400 |
| Sophie Silverstein | |
| Soomin Kim | 0000-0001-8938-2559 |
| Olga Birbrayer | |
| Niaz Banaei | 0000-0001-8501-3000 |
| Christine N. Duncan | |
| Steven P. Margossian | |
| Leslie E. Lehmann | |
| Jerome Ritz | 0000-0001-5526-4669 |
| Anthony A. Fodor | |
| Wendy B. London | 0000-0003-3571-6538 |
| Ami S. Bhatt | 0000-0001-8099-2975 |
| Jennifer S. Whangbo | |

**This file includes:**

Table S1 | Vancomycin and Polymyxin B administration details

Table S2 | Bloodstream infections

Table S3 | Shannon diversity at the species and genus taxa of patient samples at baseline (prior to any GD exposure) and two-weeks after transplant

Table S4 | Shannon diversity baseline and 2-week comparisons

Table S5 | Duration of antibiotic exposure within the first 30 days post-HCT

Table S6 | Antibiotic susceptibility testing of blood culture isolates

Table S7 | BSI Reads and Coverage based on estimated Genome size

Table S8 | Colistin (polymyxin E) resistance patterns of the Gram-negative BSIs using the disk elution test

Table S9 | Known colistin (polymyxin E) resistance genes detected in the assembly of the Gram-negative BSIs based on RGI analysis

Fig. S1 | CONSORT Flow Diagram for ClinicalTrials.gov Identifier: NCT02641236

Fig. S2 | Actual oral vancomycin-polymyxin B administration.

Fig. S3 | Samples collected and sequenced per patient.

Fig. S4 | Number of reads per sample at each pre-processing step.

Fig. S5 | Taxonomy and Shannon diversity of all samples (Genus level)

Fig. S6 | Shannon diversity is similar between the GD and no-GD groups based on intention-to-treat analysis at the genus taxonomic level.

Fig. S7 | Shannon diversity with all samples in the study.

Fig. S8 | Principal coordinates analysis (PCoA) showing most samples are not different from one another (beta diversity).

Fig. S9 | Kaplan-Meier curves of OS and PFS.

Fig. S10 | Relative abundance of gut microbes and the microbe causing the BSI in relation to antibiotic administration and Shannon diversity over time for patients C03 and C11.

Fig. S11 | Multiple BSI isolates are nearly identical or identical to those found in the gut microbiota prior to the BSI.

Fig. S12 | Antibiotic, antifungal, and antiviral administration timing

Fig. S13 | Potential model of oral vancomycin-polymyxin B on the microbiota.

Document S1 | Informal survey of pediatric HSCT centers

Document S2 | Clinical inclusion and exclusion criteria

Document S3 | CONSORT checklist

Supplementary References

**Supplemental Table 1. Vancomycin and Polymyxin B administration details**

Participants assigned to arm A received non-absorbable, oral vancomycin-polymyxin B capsules according to body surface area; approximately 250-500 mg/m2 BSA of vancomycin and approximately 125-250 mg/m2 of polymyxin B.
Each capsule contains 125 mg of vancomycin and 62.5 mg of polymyxin B.

| Body Surface Area ($m^2$) | Dose of Vancomycin-Polymyxin B Capsules |
|---|---|
| < 0.5 $m^2$ | 1 cap PO TID |
| 0.5 – 0.99 $m^2$ | 2 cap PO TID |
| 1 – 1.49 $m^2$ | 3 cap PO TID |
| ≥ 1.5 $m^2$ | 4 cap PO TID |

Vancomycin-polymyxin B were given orally or via enteric feeding tubes. Doses were repeated if the participant vomited within 30 minutes of medication administration. For administration via enteric feeding tubes, vancomycin-polymyxin B capsules may be opened and the contents dissolved in 2-3 mL bottled or sterile water at room temperature. See Supplemental Figure 2 for actual administration during the study.

**Supplemental Table 2. Bloodstream infections**

| Subject | Arm | Organism | Day of BSI episode relative to transplant |
|---|---|---|---|
| C03 | B (no-GD) | *Leclercia adecarboxylata* | 22 |
| C04 | B (no-GD) | *MRSA (Staphylococcus aureus)* *<br>*Klebsiella oxytoca*<br>*MRSA (Staphylococcus aureus)* * | 5<br>18<br>94 |
| C10 | B (no-GD) | *E. coli* | 8 |
| C11 | A (GD) | *Bacillus* by clinical microbiology lab<br><br>Sequencing results of BSI isolate:<br>*Lysinibacillus fusiformis*<br>(also called *Bacillus fusiformis*) | 31 |
| C20 | B (no-GD) | *Staphylococcus epidermidis* * | 23 |
| C22(B) | B (no-GD) | *Rothia dentocariosa*<br>*Enterococcus faecium* | 6 [#]<br>20 [#] |

* non-mucosal barrier injury (MBI) pathogens defined by the National Healthcare Safety Network (NHSN) criteria (Jan. 2021) of the Centers for Disease Control (CDC)

### BSI occurred during the second transplant; numbering is according to the date relative to the start of the second transplant

**Supplemental Table 3.**

**(A) Shannon diversity at the species level of patient samples at baseline (prior to any GD exposure) and two-weeks after transplant.**

| Species | GD (Baseline) | GD (2-week) | No-GD (Baseline) | No-GD (2-week) |
|---|---|---|---|---|
| Median | 3.55 | 2.36 | 3.31 | 3.09 |
| Range | 2.1 - 4.5 | 0.03 - 5.3 | 1.2 - 4.2 | 2.1 - 3.7 |

**(B) Shannon diversity at the genus level of patient samples at baseline (prior to any GD exposure) and two-weeks after transplant.**

| Genus | GD (Baseline) | GD (2-week) | No-GD (Baseline) | No-GD (2-week) |
|---|---|---|---|---|
| Median | 2.23 | 1.79 | 1.47 | 1.65 |
| Range | 0.6 - 3.0 | 0.05 - 4.5 | 0.3 - 3.0 | 1.2 - 2.4 |

**Supplemental Table 4. Shannon diversity baseline and 2-week comparisons.** p-value for comparing baseline samples prior to GD exposure and 2-week timepoints. Wilcoxon sign-rank for matched samples, Wilcoxon rank-sum test (Mann-Whitney *U* test) for unrelated (GD vs No-GD) comparisons

| Cohort | Comparison | *p*-value for Shannon diversity (Genus) | *p*-value for Shannon diversity (Species) |
|---|---|---|---|
| GD | Baseline vs 2-weeks post HCT | >0.99 | 0.106 |
| No-GD | Baseline vs 2-weeks post HCT | 0.922 | 0.492 |
| Overall, at Baseline | GD vs No-GD | 0.315 | 0.353 |
| Overall, at 2-weeks post HCT | GD vs No-GD | 0.796 | 0.436 |

**Supplemental Table 5. Duration of antibiotic exposure within the first 30 days post-HCT**

| | # Days of antibiotics [median (range)] | | p-value |
|---|---|---|---|
| | GD (n=10) | no-GD (n=10) | |
| CLASSES | | | |
| Cephalosporins | 0 (0, 11) | 0 (0, 8) | 0.80 |
| Fluoroquinolones | 2.5 (0, 24) | 4 (0, 17) | 0.40 |
| Broad spectrum with anaerobic coverage (Amp/Sulbactam, Pip/Tazo, Meropenem) | 13 (0, 39) | 17 (0, 32) | 0.68 |
| INDIVIDUAL ANTIBIOTICS | | | |
| Vancomycin (IV) | 0 (0, 22) | 2 (0, 19) | 0.77 |
| Meropenem | 0 (0, 19) | 0 (0, 24) | 0.69 |
| Piperacillin/Tazobactam | 7 (0, 31) | 11.5 (0, 26) | 0.49 |
| Trimethoprim-sulfamethoxazole ** | 5 (4, 5) | 5 (0, 5) | 0.23 |

** Prophylactic dose Trimethoprim / Sulfamethoxazole (TMP/SMX; cotrimoxazole) analyzed from day -5 to day +30; three patients (C04, C12 and C22) in the no-GD arm had pentamidine during the pre-treatment phase (and thus no TMP/SMX during this window). Subject C04 had prophylactic dose TMP/SMX from day 25 to 27.

**Supplemental Table 6. Antibiotic susceptibility testing of blood culture isolates.**

| Subject | BSI Organism | AMP | Oxacillin | Pip/Tazo | Amp/Sulbactam | Cefazolin (1st) | CFTX (3rd) | Cefotax. (3rd) | Ceftaz. (3rd) | Cefepime (4th) | Clindamycin | Amikacin | Gentamicin | Erythromycin | Meropenem | Ciprofloxacin | TMP/SMX | Tetracycline | Vancomycin | Linezolid | synercid (quinupristin/dalfopristin) | Tigecycline | Metronidazole | Colistin |
|---|---|---|---|---|---|---|---|---|---|---|---|---|---|---|---|---|---|---|---|---|---|---|---|---|
| C03 | *Leclercia adecarboxylata* | S | | S | S | | S | | S | S | | S | S | | S | S | S | | | | | | | I |
| C04 | *MRSA* (first infection, day +5) | | R | | R | R | R | R | | | R | | | R | | | S | S | S | S | S | S | | |
| C04 | *MRSA* (second infection, day +94) | | R | | R | R | R | R | | | R | | | R | | | S | S | S | S | S | S | | |
| C04 | *Klebsiella oxytoca_1* | | | R | R | | I | | | S | | S | S | | S | S | S | | | | | | | R |
| C04 | *Klebsiella oxytoca_2* | | | | R | | S | | | S | | S | S | | S | S | S | | | | | | | R |
| C10 | *Escherichia coli_1* | R | | R | R | | R | | R | R | | S | S | | S | R | R | | | | | | | I |
| C10 | *Escherichia coli_2* | R | | R | R | | R | | R | R | | S | S | | S | R | R | | | | | | | I |
| C11 | *Bacillus* (not anthracis, not cereus) | | S | | | | I | | | | | | S | | S | | | | S | | | | | |
| C20 | *Staph epidermidis (CoNS)* | | R | | R | R | R | R | | | R | | | R | | | R | S | S | S | S | S | | |
| C22 | *Rothia dentocariosa* (beta-lactamase negative) | | S | S | S | | | | | | | | | | S | | | | | | | | I | |
| C22 | *Enterococcus faecium* | R | R | | | | | | | | | | | | | | | | S | | | S | | |

S, susceptible; I, intermediate; R, resistant.
_1 (strain #1) _2 (strain #2) on the same day; vs (first) separate time infections (second)
Red = Gut Decontamination (GD). Blue = no-GD

A total of 9 bloodstream infection (BSI) events occurred in 6 patients (also see Figure 5 and Supplemental Figure 10). 11 clinical isolates were obtained from the clinical microbiology lab. Listed are the antibiotics along with the individual antibiotic sensitivity from the clinical microbiology laboratory of all isolates; S = susceptible, I = intermediate, R = resistant. For strains isolated on the same day, an underscore is noted (e.g., _1 = strain 1, _2 = strain 2). See Supplementary Table 8 for colistin (polymyxin E) interpretation.

Antibiotic susceptibility testing on isolates from bloodstream infections was performed by the Clinical Microbiology Laboratory at Boston Children's / Dana Farber Cancer Center, except for colistin (polymyxin E), which was performed at Stanford Health Care Clinical Microbiology Laboratory using the disk elution test as previously described [1]. Minimal inhibitory concentrations (MIC) for Enterobacteriales were interpreted using breakpoints according to Clinical and Laboratory Standards Institute (CLSI) [2] and the European Committee on Antimicrobial Susceptibility Testing (EUCAST) [3].
AMP = Ampicillin, PipTazo = Piperacillin / Tazobactam, Amp/Sulbactam = Ampicillin / Sulbactam, CFTX (3rd) = Ceftriaxone (third-generation cephalosporin), Cefotax. (3rd) = Cefotaxime (third-generation cephalosporin), Ceftaz. (3rd) = Ceftazidime (third-generation cephalosporin), Clinda = Clindamycin, Mero = Meropenem, Cipro = Ciprofloxacin, TMP/SMX = Trimethoprim / Sulfamethoxazole (cotrimoxazole), Vanc = Vancomycin

#### Supplemental Table 7. BSI Reads and Coverage based on estimated Genome size

| Sample | Raw reads | Deduplicated Reads | Trimmed Reads | Reads after host removal | Estimated size of bacterial genome (Mb) | Coverage |
|---|---|---|---|---|---|---|
| *Leclercia adecarboxylata* | 31,620,619 | 21,161,570 | 18,403,419 | 17,355,375 | 4.62 | 563x |
| *MRSA* (first infection, day +5) | 31,300,420 | 17,730,130 | 14,543,633 | 9,929,725 | 2.83 | 526x |
| *MRSA* (2nd infection, day +94) | 26,421,425 | 15,932,677 | 13,362,581 | 9,332,184 | 2.83 | 495x |
| *Klebsiella oxytoca* (Strain #1) | 43,652,689 | 28,556,138 | 24,944,940 | 23,524,208 | 6.02 | 586x |
| *Klebsiella oxytoca* (Strain #2) | 27,575,433 | 19,262,474 | 16,824,826 | 15,779,072 | 6.02 | 393x |
| *E. coli* (Strain #1) | 46,579,261 | 28,337,882 | 24,539,704 | 22,746,871 | 5.12 | 666x |
| *E. coli* (Strain #1) | 40,899,722 | 23,939,870 | 20,715,391 | 19,224,972 | 5.12 | 563x |
| *Bacillus* *(Lysinibacillus fusiformis / Bacillus fusiformis)* | 33,318,357 | 20,722,144 | 17,793,904 | 13,650,888 | 5.06 | 405x |
| *Staphylococcus epidermidis** | 45,968,718 | 25,508,418 | 22,100,194 | 20,450,659 | 2.52 | N/A* |
| *Enterococcus faecium* | 26,366,144 | 14,524,716 | 11,253,613 | 7,532,822 | 2.92 | 387x |
| *Rothia dentocariosa* | 30,518,763 | 17,751,972 | 15,060,078 | 11,092,660 | 2.49 | 668x |
| Mean | 34,929,232 | 21,220,726 | 18,140,208 | 15,510,858 | Mean | 525x |
| Median | 32,469,488 | 20,941,857 | 17,967,056 | 15,644,965 | Median | 545x |

* We were unable to do strain-level analysis as the original *Staph. epidermidis* BSI-causing isolate was archived, but upon sequencing was identified to be *E. coli*, a likely contaminant of the archived culture.

Coverage is the number of times a genome has been sequenced. Based on the Lander-Waterman equation [4] of C = LN / G

C = Coverage
L = read length
N = number of reads
G = haploid genome length

This is estimated by the number of reads after host removal over the estimated genome size. In general, we require 5-10x coverage for taxonomic identification, and 30-100x coverage for assembly of a genome (though coverage may need to be greater for highly repetitive regions, which may not be resolved with short-read sequencing).

**Supplemental Table 8. Colistin (polymyxin E) resistance patterns of the Gram-negative BSIs using the disk elution test as previously described [1]**.

| Gram-negative BSI organism | MIC | Interpretation |
|---|---|---|
| *Leclercia adecarboxylata* | <1 ug/mL | Intermediate |
| *Klebsiella oxytoca* (Strain #1) | >4 ug/mL | Resistant |
| *Klebsiella oxytoca* (Strain #2) | >4 ug/mL | Resistant |
| *E. coli* (Strain #1) | <1 ug/mL | Intermediate |
| *E. coli* (Strain #2) | <1 ug/mL | Intermediate |

Minimal inhibitory concentrations (MIC) were interpreted using breakpoints according to Clinical and Laboratory Standards Institute (CLSI) [2] and the European Committee on Antimicrobial Susceptibility Testing (EUCAST) [3]; Epidemiological cut-offs are approximately 2 ug/mL for both Klebsiella and E. coli based on EUCAST, and extrapolated to *Leclercia* based on Enterobacterales susceptibility.

**Supplemental Table 9. Known colistin (polymyxin E) resistance genes detected in the assembly of the Gram-negative BSIs based on RGI analysis [5] (strict and perfect hits only).**

| Antibiotic Resistance Gene: | *Leclercia* | *Klebsiella*_#1 | *Klebsiella* _#2 | *E. coli*_#1 | *E. coli*_#2 |
|---|---|---|---|---|---|
| PmrF | - | + | + | + | + |
| PmrA | - | - | - | - | - |
| arnA | - | - | - | - | - |
| arnT | - | + | + | - | - |
| eptA | - | - | - | + | + |
| eptB | - | - | - | + | + |
| acrA | - | - | - | + | + |
| acrB | + | + | + | + | + |
| acrR | + | - | - | + | + |
| kpnEF | + | + | + | + | + |
| MCR-9.1 | + | - | - | - | - |

\+ = Gene detected in assembled genome
\- = Gene not detected

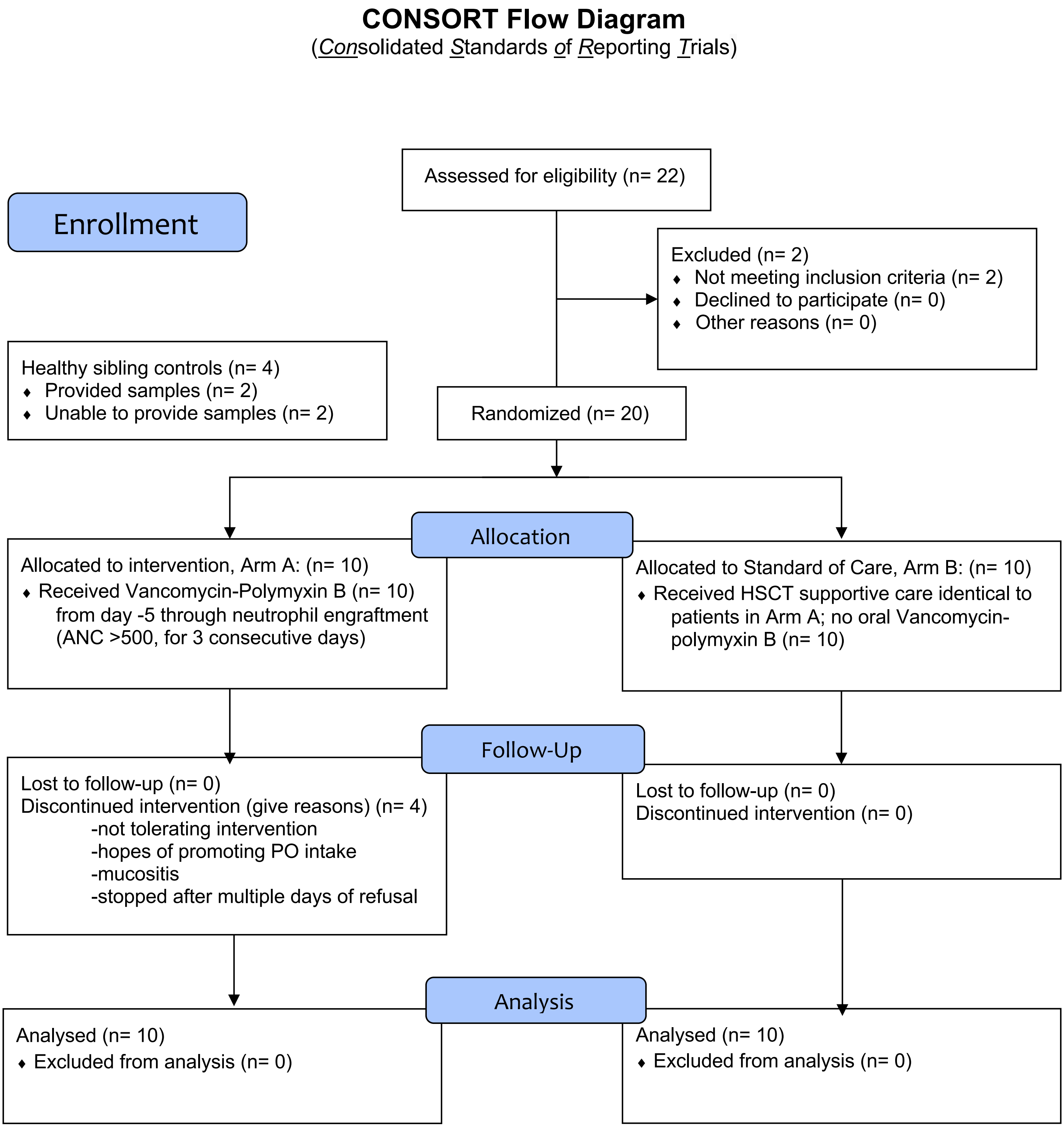


**Supplemental Fig. 1 | CONSORT Flow Diagram for ClinicalTrials.gov Identifier: NCT02641236**

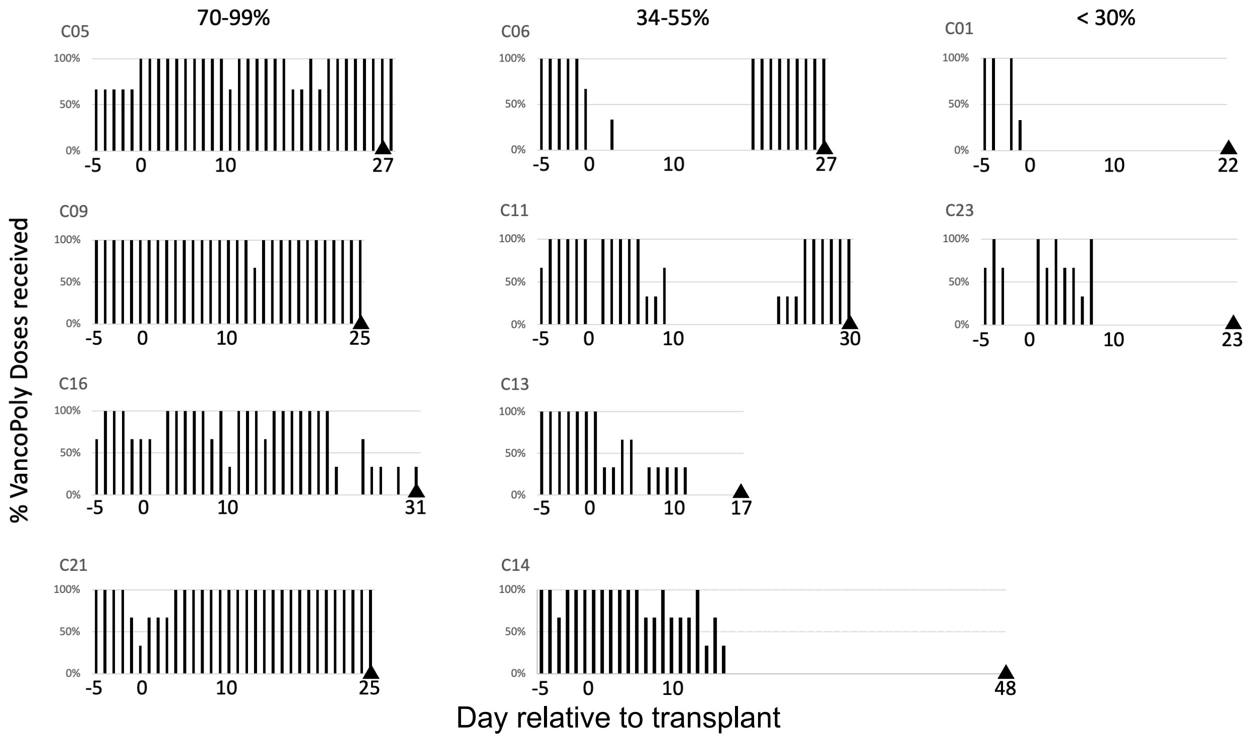


**Supplemental Fig. 2 | Actual oral vancomycin-polymyxin B administration**. Percent of doses taken daily for all 10 patients receiving gut decontamination (arm A) with oral Vancomycin-Polymyxin B starting from transplant day -5 through neutrophil engraftment (triangle) for each patient. Patients are grouped by 70-99% of doses taken (left), 34-55% (center), and <33% (right). Median day of neutrophil engraftment is day +25 for all patients, +26 for arm A only.

A.

B.

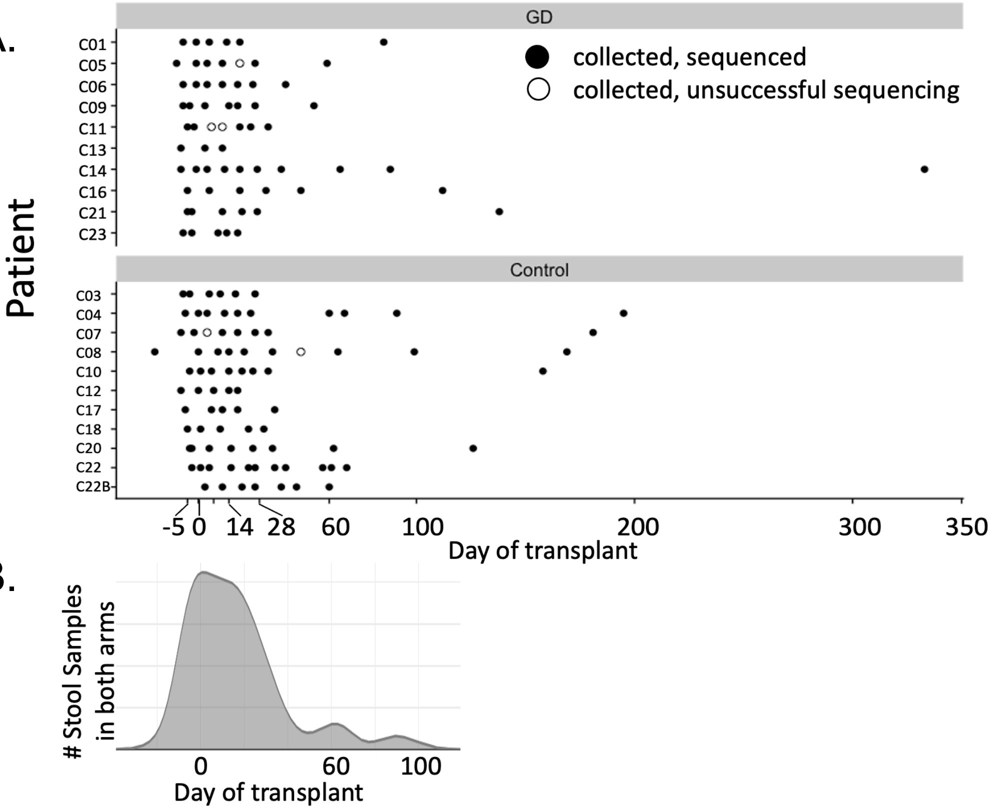


C.

**Mean, median, and range of samples collected per patient**

| | GD | No-GD | TOTAL |
|---|---|---|---|
| Mean | 6.4 | 8.3 | 7.4 |
| Median | 6.5 | 8 | 7 |
| Range | 3 to 10 | 5 to 18 | 3 to 18 |

**Supplemental Fig. 3 | Samples collected and sequenced per patient**.
(A) Stool samples collected per patient in relation to days relative to transplantation, divided into gut decontamination (GD with vancomycin-polymyxin B) and no-GD.
Closed circles are stool samples able to be sequenced, and open circles are stool samples collected but unable to be sequenced due to limited biomass. Patient 22 did not engraft and received two transplants, divided into C22 and C22B. 8 samples are greater than day 100.
(B) Total number of stool samples across all patients are focused from day -5 to the first 30 days.
(C) Table showing numbers of samples collected per patient. Total number sequenced: 143 /147 = 97% of collected samples.
18 samples total for patient 22, (divided into 1st and 2nd transplant, with 11 samples and 7 samples, respectively), and 3 samples from healthy siblings (not included in this table; Data from healthy siblings included in Supplemental Figures 5 and 8).

| Mean | Median |
|---|---|
| 63.7 x $10^6$ | 72.3 x $10^6$ |
| 41.4 x $10^6$ | 45.4 x $10^6$ |
| 35.2 x $10^6$ | 38.2 x $10^6$ |
| 16.6 x $10^6$ | 16.9 x $10^6$ |

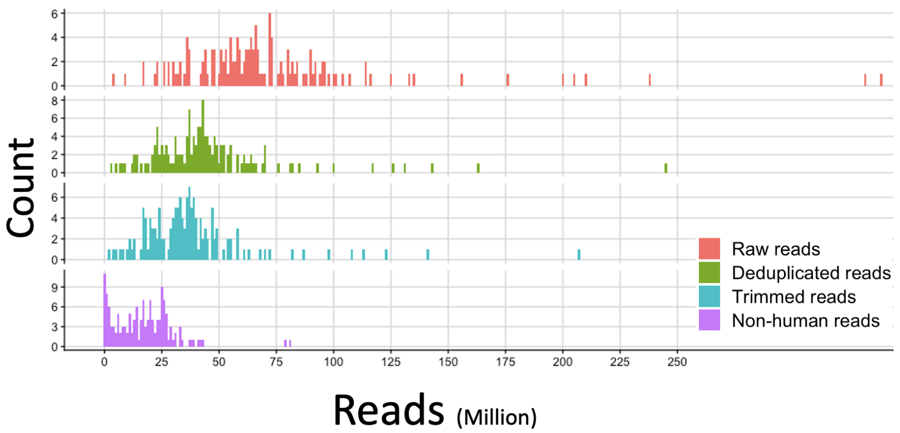


**Supplemental Fig. 4 | Number of reads per sample at each pre-processing step.**
Raw reads were demultiplexed by unique barcodes (bcl2fastq v2.20.0.422, Illumina).
Reads were deduplicated to remove PCR artifacts and any duplicates with SuperDeduper v1.4.
Deduplicated reads were trimmed using TrimGalore v0.4.4.
Finally, any human reads aligning to human genome (hg19.fa) were removed prior to analysis.
Two samples were mostly human reads and had <100,000 microbial reads after removing human reads, however these samples had 21.3 and 48.8 x $10^6$ reads after deduplication and trimming.
The final mean read-depth was 16.6 Million reads per sample.

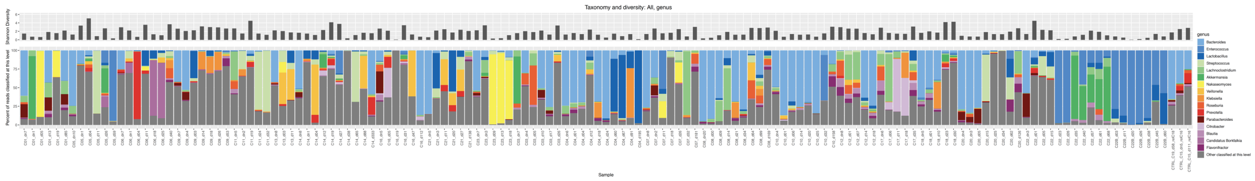


**Supplemental Fig. 5 |** Taxonomy of all samples displayed as the relative abundance as a percentage of classified reads at the genus level.  Shannon diversity at each time point is shown as a bar.

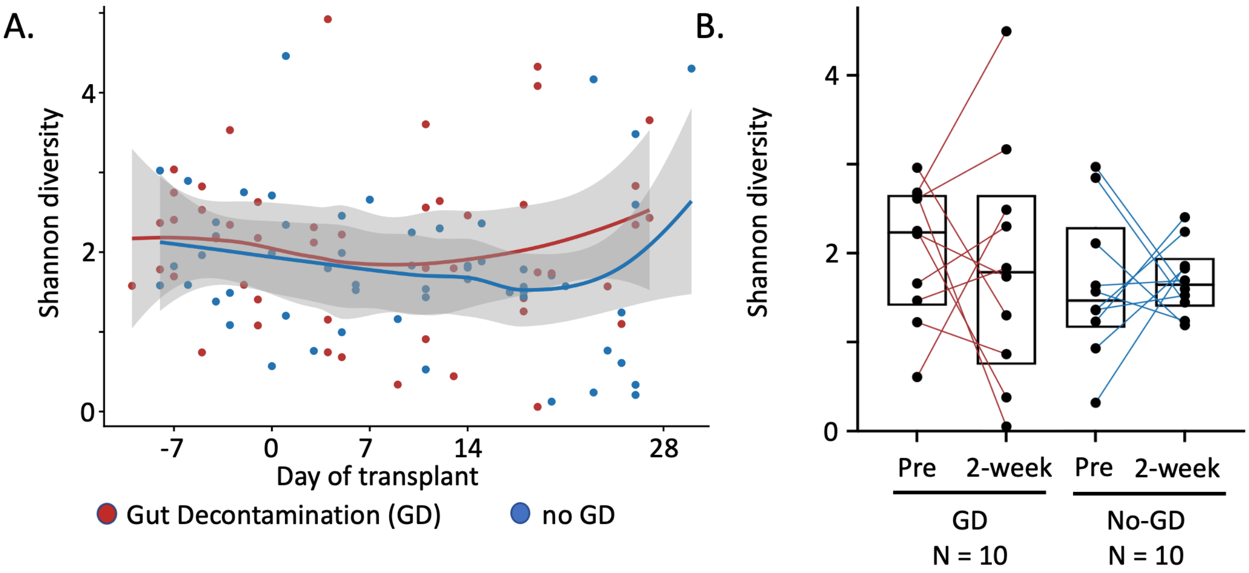


**Supplemental Figure 6 | Shannon diversity is similar between the GD and no-GD groups based on intention-to-treat analysis at the genus taxonomic level**. Red are samples from patients undergoing GD and blue is no-GD arm.

(A) Shannon diversity over time analyzed at the genus level using local polynomial regression fitting (LOESS-locally estimated scatterplot smoothing of the mean Shannon diversity) showing similarity between the two groups.

(B) Shannon diversity of individual patient data from pre-transplant (before GD antibiotics) to 2 weeks post HCT connected with a line. Boxes shown are the median with hinges at the 25% and 75%. All comparisons not significant (see Supplemental Table 4).

N=10 subjects GD arm, N=10 subjects no-GD arm.

Comparison at the species level (see Figure 2 and Suppl. Table 4) is also not significant.

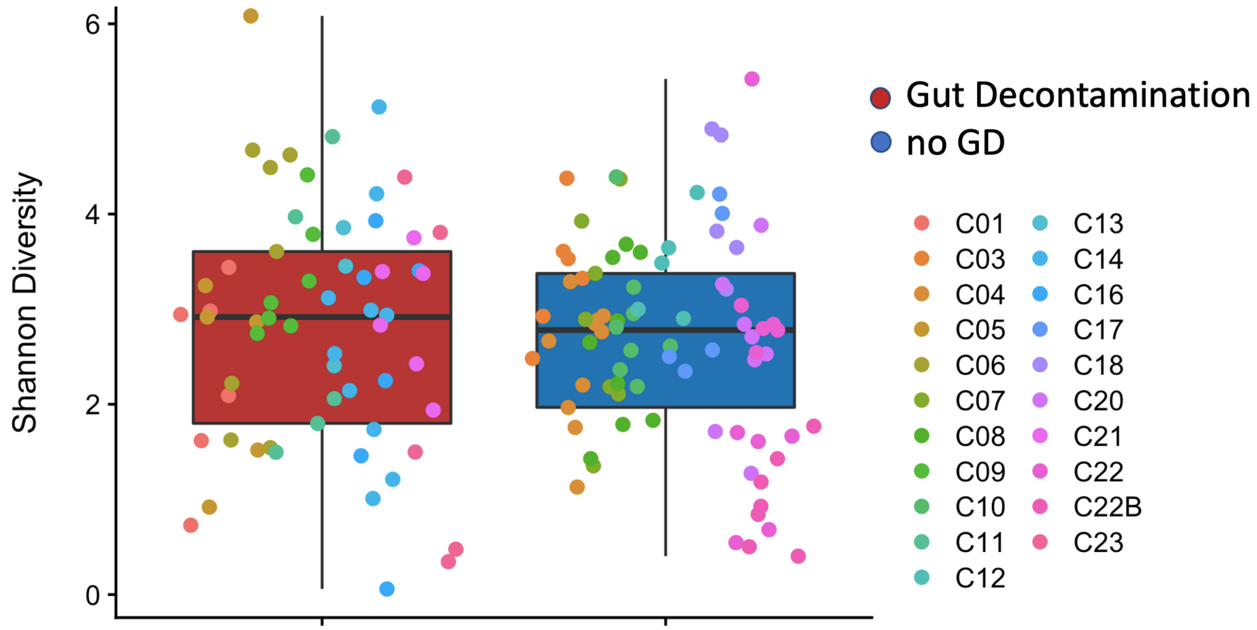


**Supplemental Fig. 7 | Shannon diversity with all samples in the study**.
Box and whiskers plot of the mean Shannon diversity index shown at the species level with the lower and upper hinges correspond to the first and third quartiles (the 25th and 75th percentiles). Boxes are labeled as gut decontamination (red) and no-GD (blue). Whisker extends from the hinge to the largest value no further than 1.5 * inter-quartile range (IQR) from the hinge. Individual patient samples are colored dots according to the legend. Statistical test via Wilcoxon with FDR, $p_{adj.} = 0.51$. Subject C22 divided into C22 (first transplant) and C22B (second transplant) for clarity of diversity trends between transplants.

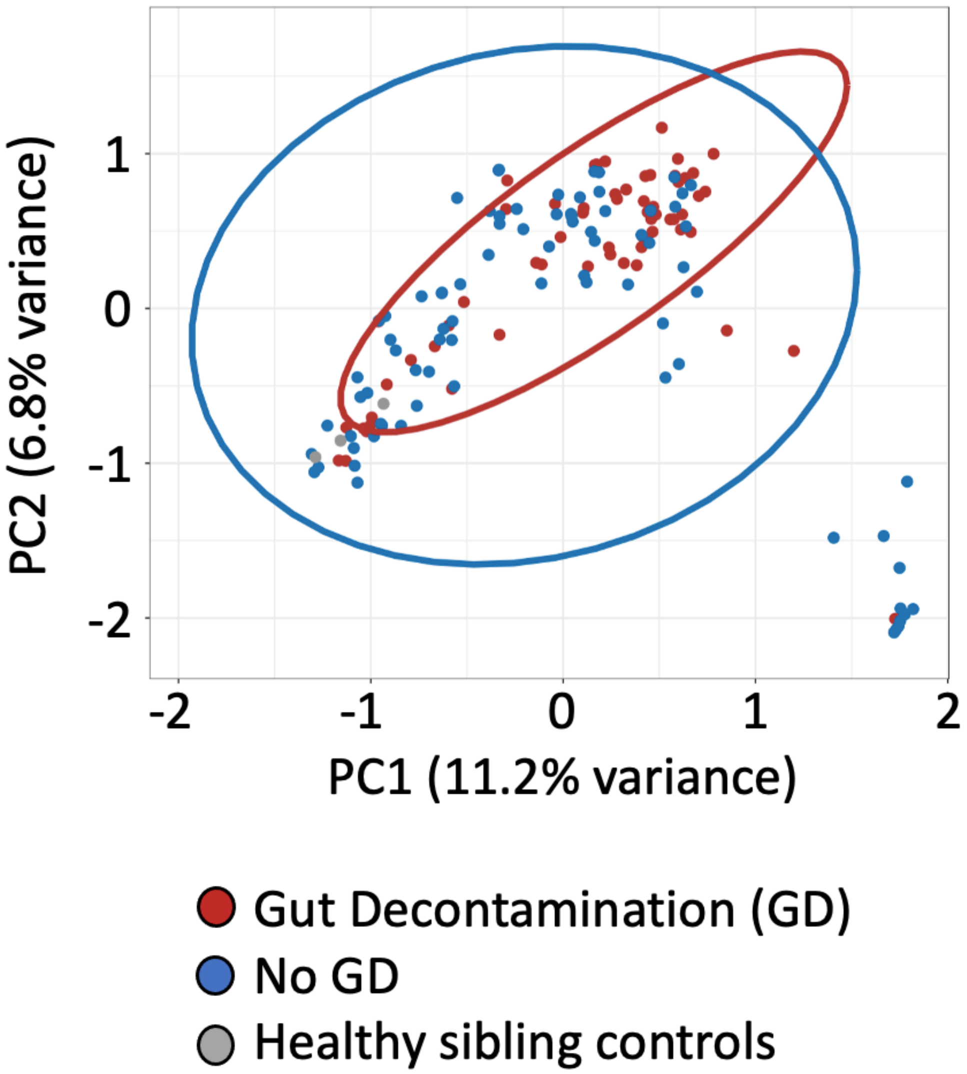


**Supplemental Figure 8 | Principal coordinates analysis (PCoA) showing most samples are not different from one another (beta diversity)**.

Samples from patients undergoing GD (red), no-GD (blue), and three samples from two healthy sibling controls (gray).

PCoA using a Bray-Curtis dissimilarity index at species level indicates that 11.2% and 6.8% of the variance can be explained by the first two principal coordinates. Ellipses represent the 95% CI for the GD and no-GD groups. A group of outliers (from 3 different patients, C05, C07, and C22) in the lower right corner include any sample with >45% relative abundance of *Enterococcus faecium.*

As a control, stool samples from two healthy sibling donors were also collected to serve as a comparison to the HCT patients, and were similar at the time points collected (all in the lower left corner). Between Control Sibling C15 and Patient C16 (GD arm), the Analysis of Similarity (ANOSIM) statistic R was -0.3125 with a significance of 0.928 after 999 permutations. Between Control Sibling C19 and Patient C18 (no-GD arm) the ANOSIM statistic R was -0.04 with a significance of 0.5 after 999 permutations.

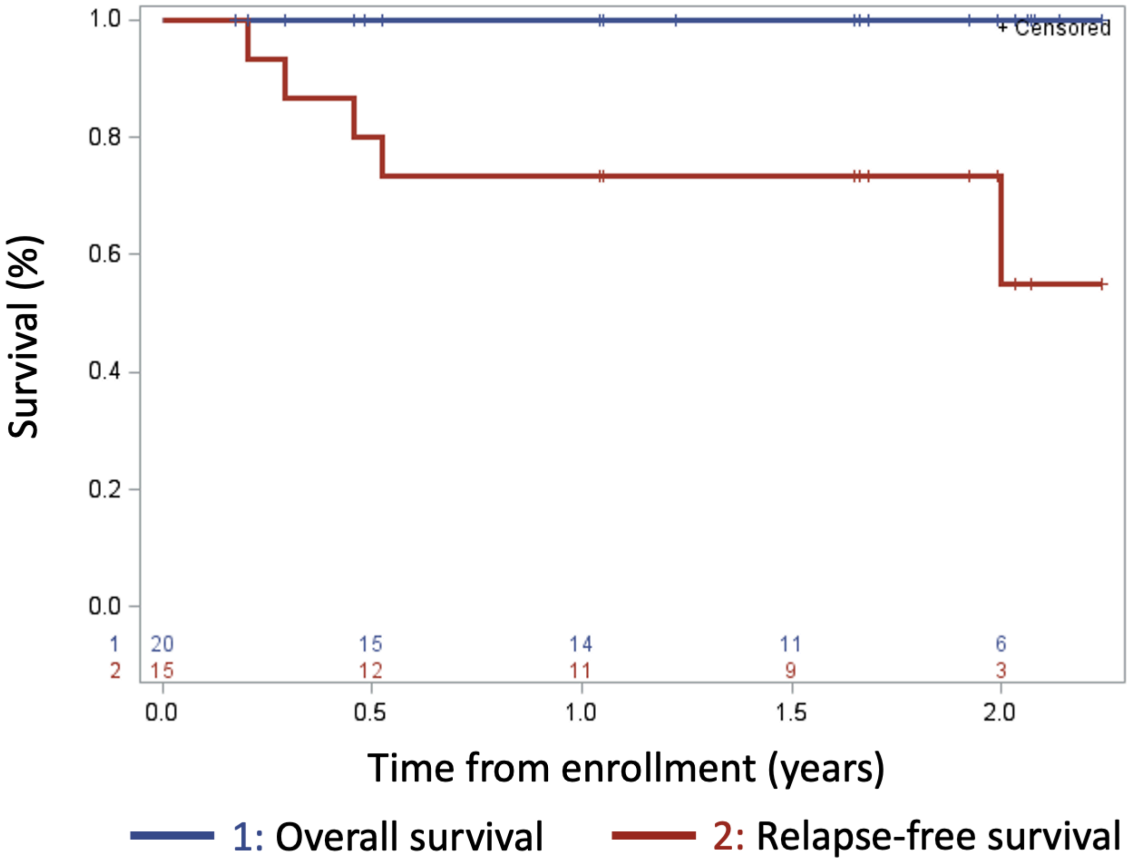


**Supplemental Figure 9 | Kaplan-Meier curves of OS (n=20 overall) and relapse-free survival (n=15 with malignancy)**

For OS at 1-year, no patients died; the 1-year OS was 100% (n=20). For relapse-free survival, of the 15 patients with a malignancy, five patients had a relapse; the 1-year relapse-free survival was 73±11.4% (n=15)

**A.**

C03

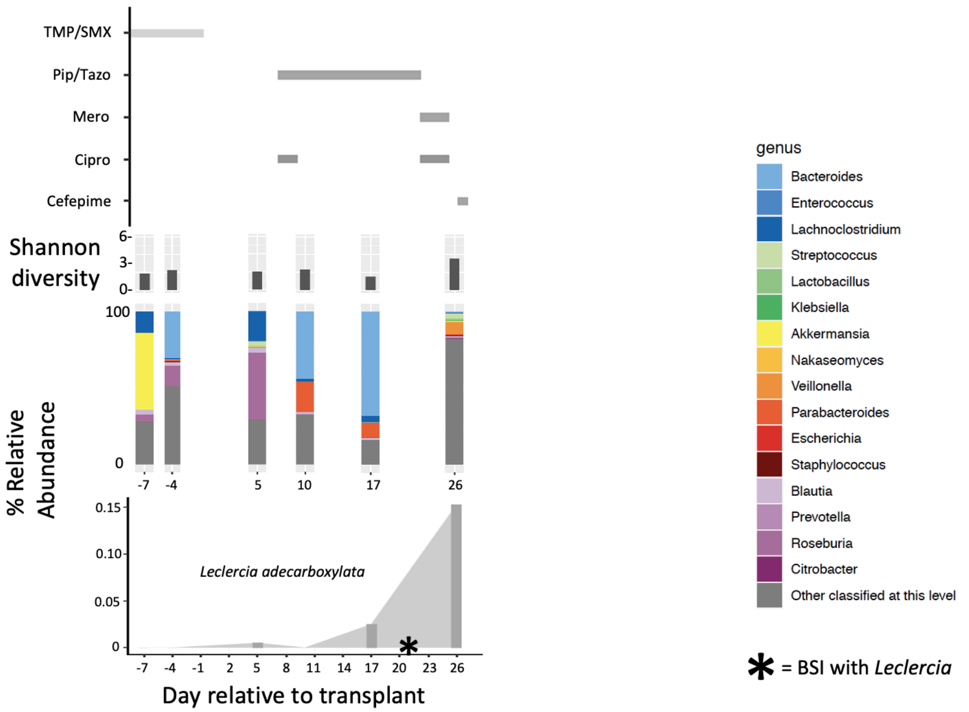


**Supplemental Fig. 10 | Relative abundance of gut microbes and the microbe causing the BSI in relation to antibiotic administration and Shannon diversity over time for patients C03 and C11.**

(A). Results for patient C03
Days relative to the course of the transplant on the X-axis, (from top to bottom on the Y-axis) antibiotic administration, alpha (Shannon) diversity, relative abundance of microbes in the stool samples at the genus taxonomic level (with organisms listed by color according to the key at the right), and relative abundance of in the gut of the BSI-causing organism with the date of the BSI shown as an asterisk (*).

**B.**

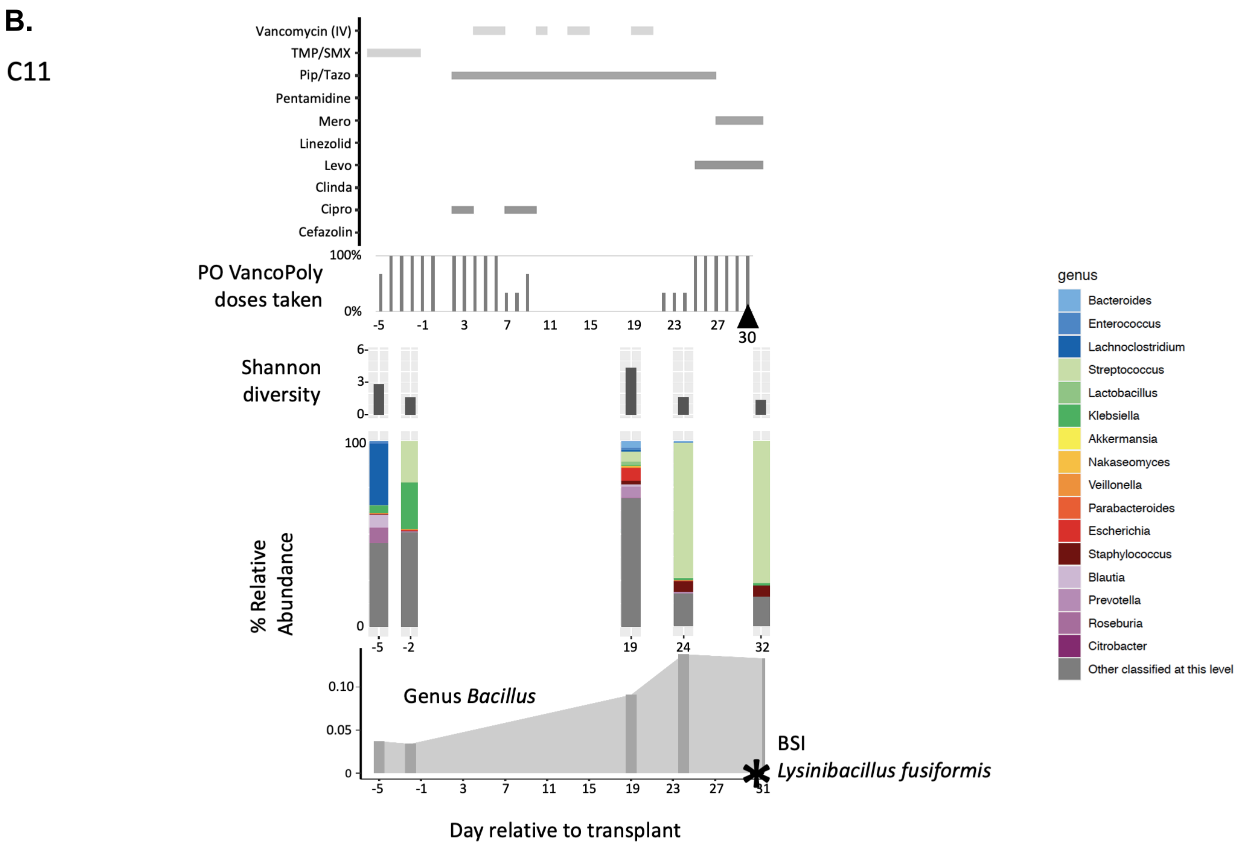


(B). Results for patient C11
Days relative to the course of the transplant on the X-axis, (from top to bottom on the Y-axis) antibiotic administration, percentage of the oral (PO) vancomycin-polymyxin b doses taken from day -5 through engraftment, alpha (Shannon) diversity, relative abundance of microbes in the stool samples at the genus taxonomic level (with organisms listed by color according to the key at the right), and relative abundance of Bacillus genus over time. Note that the BSI causing organism, *Lysinibacillus fusiformis*, was not found in any of the stool samples for this patient suggesting the BSI did not originate from the gut. Date of *Lysinibacillus fusiformis* BSI shown as an asterisk (*).

Cipro = Ciprofloxacin, Clinda = Clindamycin, Levo = Levofloxacin, Mero = Meropenem, PipTazo = Piperacillin / Tazobactam, TMP/SMX = Trimethoprim / Sulfamethoxazole (cotrimoxazole)

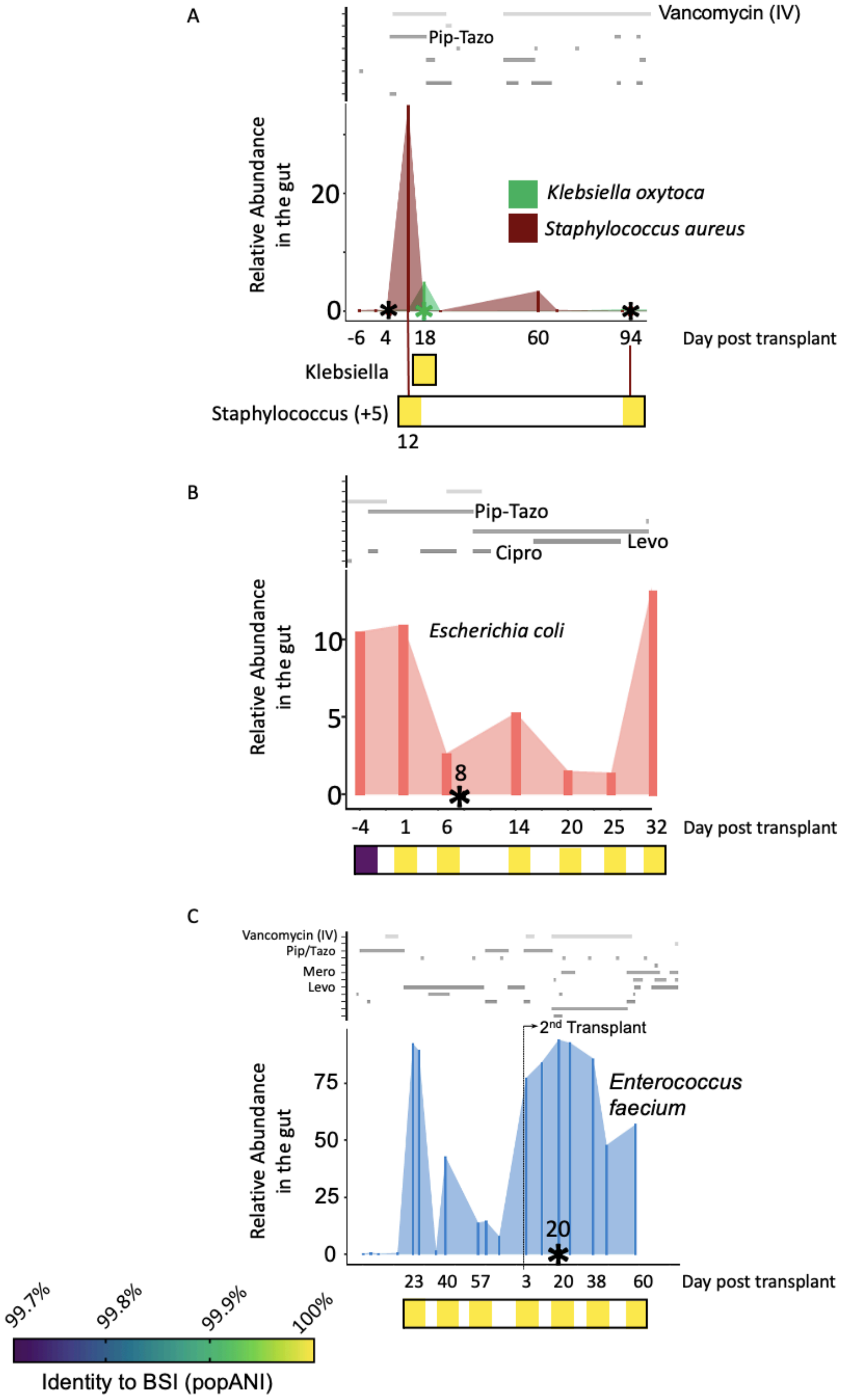


**Supplemental Fig. 11 | Multiple BSI isolates are nearly identical or identical to those found in the gut microbiota prior to the BSI**.

5 BSIs from 3 patients had sufficient coverage to be evaluated by *inStrain*. Relative abundance of the species in the gut that corresponds to the BSI. Above is the administration of antimicrobials, with select

antibiotics labeled. Below is the identity of the stool sample to the BSI measures as a population average nucleotide identity (popANI) at the time relative to the transplant date. The relative scale of the popANI is shown in the lower left corner.

A. *Klebsiella* BSI on day +18 has 100% popANI compared to the BSI on the same day. *MRSA* BSI on days +5 and +94. The stool sample from day +12 had *S. aureus* at 35% relative abundance that was identical to the BSI on day +5 (100% popANI, zero SNPs detected in 2.8 Mb of sequence compared) and nearly identical (99.9999% popANI) to BSI on day +94.

B. *E. coli* BSI on day +8 is identical (100% popANI, 5 Mb compared) to the six stool samples collected from days +1 to +32. A distinct strain was present in the stool at day -4 (99.7% popANI to BSI and other stool samples).

C. *E. faecium* BSI on day +20. *E. faecium* stool samples from day +23 of the first transplant through the end of the study (14 samples total) are nearly identical (>99.9999% popANI) to the BSI strain on day +20

See Table 3 for concordance values (coverage, popANI, population SNPs, and conANI) of BSI isolate to stool metagenomes.

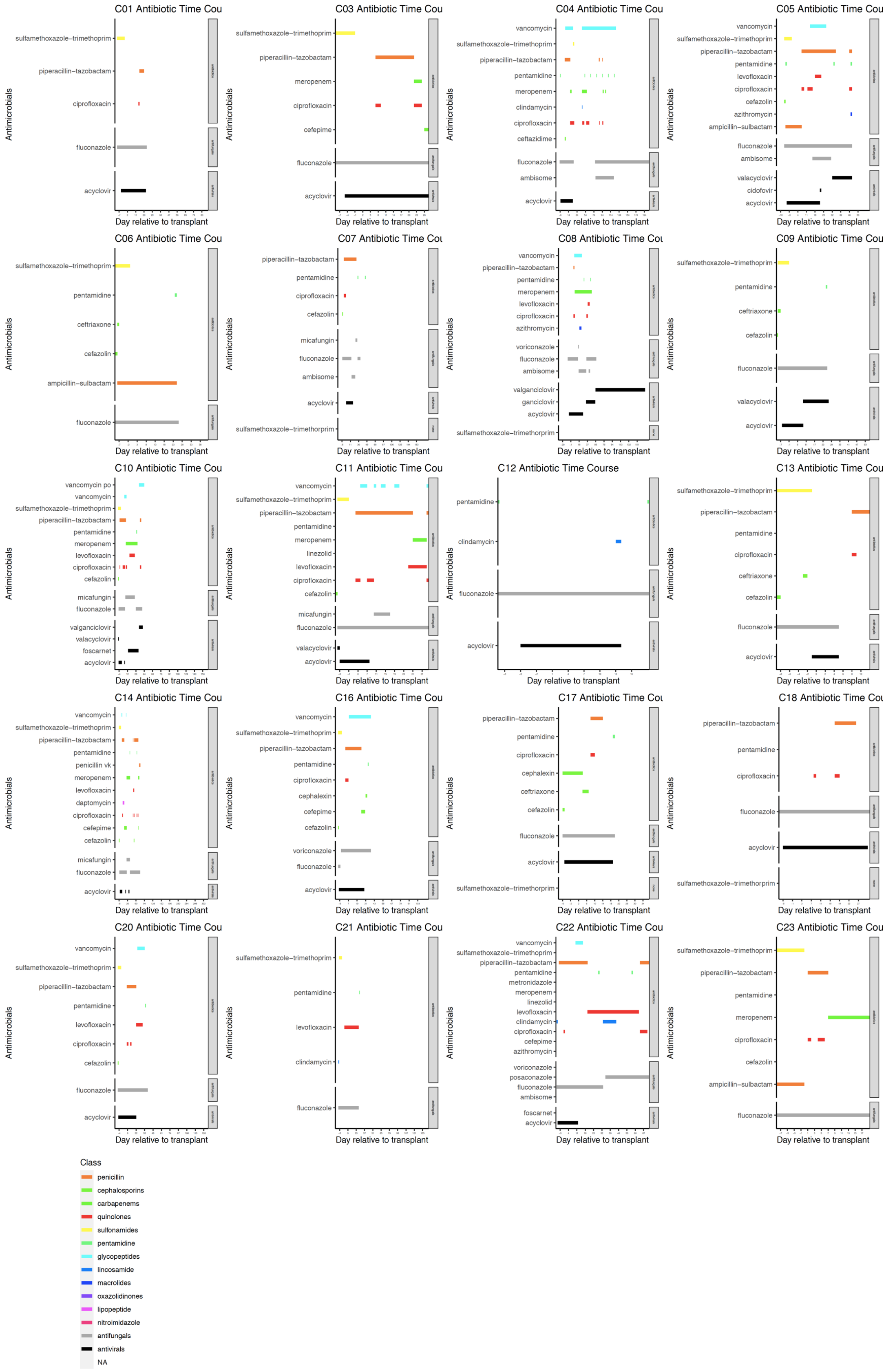


**Supplementary Fig. 12 | Antibiotic, antifungal, and antiviral administration timing**

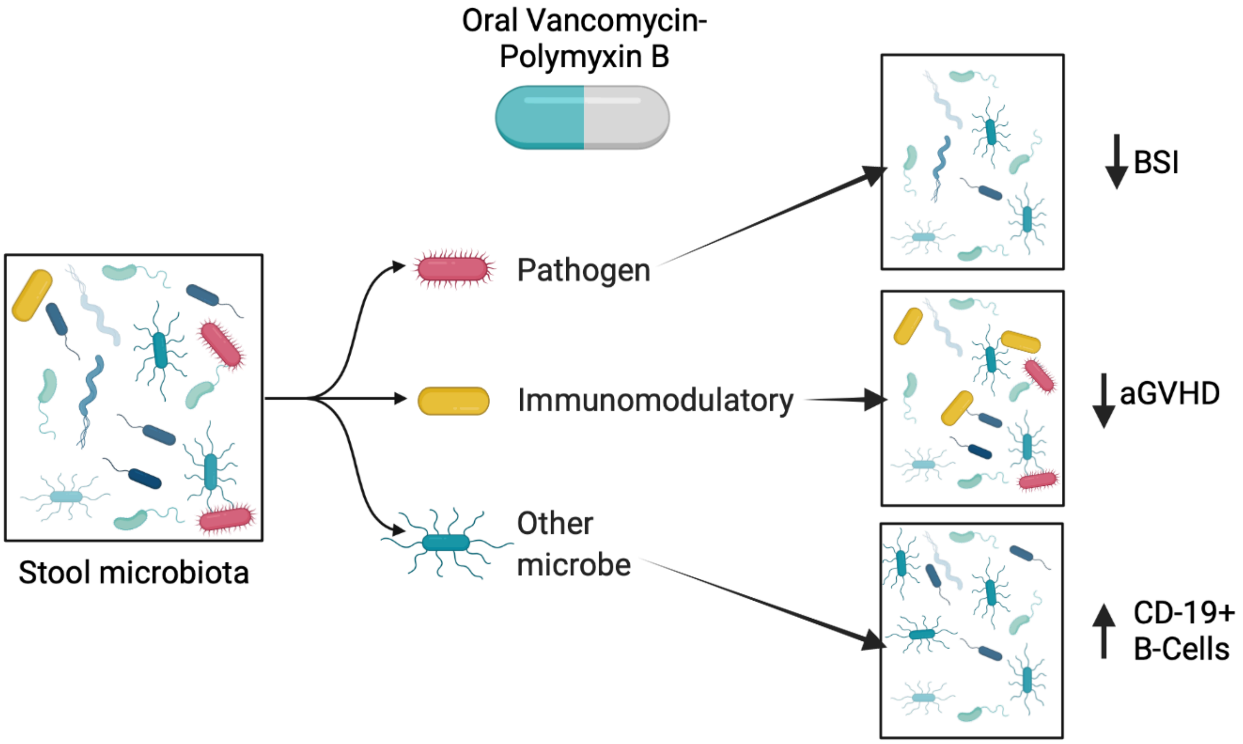


**Supplementary Fig. 13 | Potential model of oral vancomycin-polymyxin B on the microbiota**. Possible scenarios for presumed beneficial effects of oral vancomycin-polymyxin B gut decontamination on the microbiota. GD may decrease the number of pathogens in the gut that cause a BSI, may allow immunomodulatory bacteria to increase in number or create an environment where the immune system is less prone to the inflammatory aGVHD state, or allow expansion of other microbes that alter the host-microbiota interaction (such as changes in the number of CD-19+ B-cells).

#### Supplemental document 1 | Informal survey of pediatric HCT centers

| Q1: Approximately how many pediatric allogeneic hematopoietic stem cell transplants does your center perform on an annual basis? | | |
|---|---|---|
| **Prompt** | **Count** | **Pct** |
| Less than 10 per year | 6 | 19.35% |
| **10-25 per year** | **12** | **38.71%** |
| 26-35 per year | 4 | 12.90% |
| 36-50 per year | 6 | 19.35% |
| Greater than 50 per year | 3 | 9.68% |
| **Aggregates** | | |
| Count | | 31 |

| Q2: What is your estimated incidence of acute graft-versus-host disease (GVHD) among pediatric patients at your center for recipients of MATCHED SIBLING donor stem cell transplants? | | |
|---|---|---|
| **Prompt** | **Count** | **Pct** |
| Less than 5% | 4 | 13.33% |
| 5-10% | 8 | 26.67% |
| 11-15% | 7 | 23.33% |
| **16-20%** | **9** | **30.00%** |
| Great than 20% | 2 | 6.67% |
| **Aggregates** | | |
| Count | | 30 |

| Q3: What is your estimated incidence of acute graft-versus-host disease (GVHD) among pediatric patients at your center for recipients of UNRELATED donor stem cell transplants? | | |
|---|---|---|
| **Prompt** | **Count** | **Pct** |
| Less than 5% | 2 | 6.45% |
| 5-10% | 2 | 6.45% |
| 11-15% | 3 | 9.68% |
| 16-20% | 9 | 29.03% |
| **Great than 20%** | **15** | **48.39%** |
| **Aggregates** | | |
| Count | | 31 |

| Q4: Does your stem cell transplant program practice gut decontamination for acute GVHD prophylaxis? |
|---|

| Prompt | Count | Pct |
|---|---|---|
| Yes | 3 | 9.38% |
| **No** | **29** | **90.63%** |

| Aggregates | |
|---|---|
| Count | 32 |

**Supplemental document 2 | Clinical inclusion and exclusion criteria**

Inclusion Criteria:

- Eligibility Criteria for Patients Undergoing Allogeneic HSCT
  - Recipient of 9/10 or 10/10 (HLA-A, -B, -C, -DRB1, -DQB1) matched bone marrow allogeneic hematopoietic stem cell transplantation (HSCT) OR 4/6, 5/6 and 6/6 (HLA-A, -B, -DR) matched cord blood allogeneic HSCT.
  - Participants may have underlying malignant or non-malignant hematologic disease, except for primary immunodeficiency, as the indication for their allogeneic HSCT. Patients with immune dysregulation such as familial or secondary hemophagocytic lymphohistiocytosis (HLH) are eligible.
  - Participants may receive either a myeloablative or non-myeloablative(reduced-intensity) conditioning regimen. Anti-thymocyte globulin (ATG) in the conditioning regimen is permitted.
  - Graft-versus-host disease (GVHD) prophylaxis with any of the following agents: calcineurin inhibitor, and short-course methotrexate, with or without steroids, mycophenolate mofetil, and sirolimus.
  - Age ≥ 4 years old and toilet-trained. Participants must be able to deposit stool samples directly into stool collection containers. Stool specimens from diapers are difficult to obtain and are prone to more sampling error, particularly for loose or liquid stools which are common in the peri-transplant period.
  - Lansky/Karnofsky performance status ≥60%
  - Ability to understand and/or the willingness of their parent or legally authorized representative to sign a written informed consent document
- Eligibility Criteria for Healthy Bone Marrow Donors
  - Healthy individuals, ages ≥ 4 years and toilet-trained, who have been identified by BCH or DFCI providers as 9/10 or 10/10 (HLA-A, -B, -C, -DRB1, -DQB1 matched bone marrow donors for transplantation will also be eligible to participate in this study.

Exclusion Criteria:

- Patients undergoing allogeneic HSCT for correction of a primary immunodeficiency disorder (e.g. SCID).
- Patients with age ≤ 10 years undergoing HSCT with a matched sibling donor. These patients are at very low risk of acute GVHD and do not receive gut decontamination per our institutional standard practice.
- Participants receiving GVHD prophylaxis with drugs other than calcineurin inhibitors, methotrexate or steroids, and agents listed above (e.g. abatacept).
- History of allergic reactions attributed to oral vancomycin or oral polymyxin B.
- Participants undergoing active therapy for immune-mediated or infectious colitis upon admission for allogeneic HSCT.
- Participants receiving antibiotic therapy for treatment of a bacterial infection or bacterial prophylaxis upon admission for allogeneic HSCT. Use of any agent (e.g. sulfamethoxazole/trimethoprim) for prophylaxis of *Pneumocystis jirovecii* pneumonia is permitted. Concurrent use of anti-fungal and anti-viral therapies is also permitted.

- Uncontrolled intercurrent illness including, but not limited to, ongoing or active infection or psychiatric illness/social situations that would limit compliance with study requirements.

**Supplemental document 3 |** CONSORT (*Con*solidated *S*tandards *o*f *R*eporting *T*rials) checklist

| Section/Topic | Item No | Checklist item | Reported on page No. |
|---|---|---|---|
| **Title and abstract** | | | |
| | 1a | Identification as a randomized trial in the title or abstract | 1 (Title) 5 (abstract) |
| | 1b | Structured summary of trial design, methods, results, and conclusions (for specific guidance see CONSORT for abstracts) | 28-29 |
| **Introduction** | | | |
| Background and objectives | 2a | Scientific background and explanation of rationale | 5, 10-11 |
| | 2b | Specific objectives or hypotheses | 28-29 |
| **Methods** | | | |
| Trial design | 3a | Description of trial design (such as parallel, factorial) including allocation ratio | 28-29 |
| | 3b | Important changes to methods after trial commencement (such as eligibility criteria), with reasons | none |
| Participants | 4a | Eligibility criteria for participants | 28, 34, Supp. Document 2 |
| | 4b | Settings and locations where the data were collected | 34 |
| Interventions | 5 | The interventions for each group with sufficient details to allow replication, including how and when they were actually administered | 28-29, Supp. Table 1 |
| Outcomes | 6a | Completely defined pre-specified primary and secondary outcome measures, including how and when they were assessed | 28-29 |
| | 6b | Any changes to trial outcomes after the trial commenced, with reasons | none |
| Sample size | 7a | How sample size was determined | 28 |
| | 7b | When applicable, explanation of any interim analyses and stopping guidelines | N/A |
| Randomization: | | | |
| Sequence generation | 8a | Method used to generate the random allocation sequence | 28 |
| | 8b | Type of randomization; details of any restriction (such as blocking and block size) | 28 |
| Allocation concealment mechanism | 9 | Mechanism used to implement the random allocation sequence (such as sequentially numbered containers), describing any steps taken to conceal the sequence until interventions were assigned | N/A |
| Implementation | 10 | Who generated the random allocation sequence, who enrolled participants, and who assigned participants to interventions | 28 |
| Blinding | 11a | If done, who was blinded after assignment to interventions (for example, participants, care providers, those assessing outcomes) and how | N/A |

| | | | |
|---|---|---|---|
| | 11b | If relevant, description of the similarity of interventions | 28 |
| Statistical methods | 12a | Statistical methods used to compare groups for primary and secondary outcomes | 28, 33-34 |
| | 12b | Methods for additional analyses, such as subgroup analyses and adjusted analyses | 28, 33-34 |
| **Results** | | | |
| Participant flow (a diagram is strongly recommended) | 13a | For each group, the numbers of participants who were randomly assigned, received intended treatment, and were analyzed for the primary outcome | Suppl. Fig 1 |
| | 13b | For each group, losses and exclusions after randomization, together with reasons | Suppl. Fig 1 |
| Recruitment | 14a | Dates defining the periods of recruitment and follow-up | 28, 34 |
| | 14b | Why the trial ended or was stopped | N/A |
| Baseline data | 15 | A table showing baseline demographic and clinical characteristics for each group | Table 1 |
| Numbers analyzed | 16 | For each group, number of participants (denominator) included in each analysis and whether the analysis was by original assigned groups | 28, Suppl. Fig 1 |
| Outcomes and estimation | 17a | For each primary and secondary outcome, results for each group, and the estimated effect size and its precision (such as 95% confidence interval) | Figures 2, 3 Table 2 |
| | 17b | For binary outcomes, presentation of both absolute and relative effect sizes is recommended | Table 2 |
| Ancillary analyses | 18 | Results of any other analyses performed, including subgroup analyses and adjusted analyses, distinguishing pre-specified from exploratory | 28-29 |
| Harms | 19 | All important harms or unintended effects in each group (for specific guidance see CONSORT for harms) | 15 |
| **Discussion** | | | |
| Limitations | 20 | Trial limitations, addressing sources of potential bias, imprecision, and, if relevant, multiplicity of analyses | 25-26 |
| Generalizability | 21 | Generalizability (external validity, applicability) of the trial findings | 26-27 |
| Interpretation | 22 | Interpretation consistent with results, balancing benefits and harms, and considering other relevant evidence | 22-27 |
| **Other information** | | | |
| Registration | 23 | Registration number and name of trial registry | 6, 34 |
| Protocol | 24 | Where the full trial protocol can be accessed, if available | 34 |
| Funding | 25 | Sources of funding and other support (such as supply of drugs), role of funders | 6, 36 |
